# Radiomics analysis to predict pulmonary nodule malignancy using machine learning approaches

**DOI:** 10.1101/2022.10.03.22280659

**Authors:** Matthew T. Warkentin, Hamad Al-Sawaihey, Stephen Lam, Geoffrey Liu, Brenda Diergaarde, Jian-Min Yuan, David O. Wilson, Martin C. Tammemägi, Sukhinder Atkar-Khattra, Benjamin Grant, Yonathan Brhane, Elham Khodayari-Moez, Kieran R. Campbell, Rayjean J. Hung

## Abstract

**Purpose:** Screening with low-dose computed tomography can reduce lung cancer-related mortality. However, most screen-detected pulmonary abnormalities do not develop into cancer and it remains challenging to identify high-risk nodules among those with indeterminate appearance. We aim to develop and validate prediction models to discriminate between benign and malignant pulmonary lesions based on radiological features.

**Methods:** Using four international lung cancer screening studies, we extracted 2,060 radiomic features for each of 16,797 nodules among 6,865 participants. After filtering out redundant and low-quality radiomic features, 642 radiomic and 9 epidemiologic features remained for model development. We used cross-validation and grid search to assess three machine learning models (XGBoost, Random Forest, LASSO) for their ability to accurately predict risk of malignancy for pulmonary nodules. We fit the top-performing ML model in the full training set. We report model performance based on the area under the curve (AUC) and calibration metrics in the held-out test set.

**Results:** The ML models that yielded the best predictive performance in cross-validation were XGBoost and LASSO, and among these models, LASSO had superior model calibration, which we considered to be the optimal model. We fit the final LASSO model based on the optimized hyperparameter from cross-validation. Our radiomics model was both well-calibrated and had a test-set AUC of 0.930 (95% CI: 0.901-0.957) and out-performed the established Brock model (AUC=0.868, 95% CI: 0.847-0.888) for nodule assessment.

**Conclusion:** We developed highly-accurate machine learning models based on radiomic and epidemiologic features from four international lung cancer screening studies that may be suitable for assessing suspicious, but indeterminate, screen-detected pulmonary nodules for risk of malignancy.

## Introduction

Lung cancer is the leading cause of cancer mortality globally^1^. Only 10-20% lung cancer patients live up to five years after diagnosis^2^. However, several large randomized screening trials have demonstrated that low-dose computed tomography (CT) screening can significantly reduce lung cancer mortality through early detection^3–6^. The National Lung Screening Trial (NLST) observed a 20% reduction in lung cancer-related mortality following CT screening^4^, while the Dutch-Belgian trial (NELSON) observed a reduction in mortality of 24% in men and 33% in women^3^.

Despite the promise of screening, the clinical management of screen-detected pulmonary nodules and the false-positive rate are important determinants for screening program efficacy. Across several studies, the average nodule detection rate was 20%, meanwhile, more than 90% of screen-detected nodules were benign^7^. Inaccurate assessment of indeterminate nodules may lead to unnecessary diagnostic workup, including diagnostic screens (which confer higher radiation dosing), invasive procedures such as bronchoscopy, biopsy, or surgery, and may lead to overdiagnosis of indolent cancers^7^. Excess follow-up carries significant healthcare costs, utilizes critical hospital and human resources, and may lead to adverse events and complications, including death, and can cause anxiety and decreased quality of life for the screened participant.

Several guidelines have been developed to help inform indeterminate nodule management, however, there remains significant heterogeneity in these recommendations^8–^ To address these issues, probability models have been developed to help identify high-risk lesions and guide clinical decision-making^23–25^. These models have traditionally been based on patient characteristics (e.g., age, smoking history, etc.) and clinically-collected nodule morphology and textural features (e.g., size, attenuation, etc.). These features characterize important aspects of the nodule and are routinely collected as part of the clinical management of pulmonary findings.

Nodule probability models based on routinely-collected patient and nodule information have shown good performance, however, there is growing interest in leveraging medical images directly to perform automated quantitative image analysis, enabling the quantification of hundreds or thousands of radiomic features that may capture important information otherwise imperceptible to the human eye. Radiomic features quantify aspects of the 3-dimensional (3D) morphology and grayscale distribution for a region-of-interest^26^. It is expected that radiomic features, in combination with patient-level information, will be able to accurately discriminate between benign and malignant pulmonary nodules beyond what has been achieved with traditional clinical features. However, it is currently unknown which features will be most important and whether they will generalize well to other screening populations.

The goal of the current study is to perform quantitative image analysis and evaluate the predictive performance of high-dimensional radiomic features for pulmonary nodule malignancy assessment, and to develop and validate models using data from several large independent international lung cancer screening studies.

## Methods

### Lung Cancer Screening Studies

As part of the Integrative Analysis of Lung Cancer Etiology and Risk (INTEGRAL) program, we used data collected by four independent lung cancer screening studies for this analysis: 1. National Lung Screening Trial (NLST), 2. PanCanadian Early Detection of Lung Cancer (PanCan) Study, 3. International Early Lung Cancer Action Program (IELCAP-Toronto), and 4. Pittsburgh Lung Screening Study (PLuSS). Details of each study have been described previously^4, 5, 27–30^. We provide brief descriptions of each study in the following sections. Details about the study protocol used by each study are included in the Supplemental Materials.

### National Lung Screening Trial (NLST)

NLST was a large randomized multi-center lung cancer screening study comparing low-dose helical CT to standard chest radiography (CXR) for screening adult heavy smokers^4, 5^. Eligible participants were age 55 to 74 years, with 30 or more pack-years history of smoking, and former smokers quitting no more than 15 years prior. NLST enrolled 53,456 participants across 33 centers in the United States in 2002. We only use image data from the CT screening arm in the current study. Positive screen-detected findings were considered as any non-calcified nodules (NCN) with a diameter of 4mm or greater.

### Pan-Canadian Early Detection of Lung Cancer Study (PanCan)

PanCan was a multi-center, single-arm prospective lung cancer screening study that included 2,537 participants^27^. Participants were recruited from eight sites across Canada. Eligible participants included those 50 to 75 years of age, without a self-reported history of lung cancer, current or former smokers, an estimated 6-year risk of lung cancer of at least 2% based on an earlier edition of the PLCOm2012 model^31^, and an ECOG performance status of 0 or 1. Screening was performed with multi-detector row CT scanners. Each scan was reviewed by a train radiologist and up to 10 lung nodules were identified and recorded.

### International Early Lung Cancer Action Program (IELCAP-Toronto)

IELCAP was an international single-arm multi-centre study evaluating low-dose CT for lung cancer screening of high-risk individuals^28, 29^. A common study protocol was adopted for screening regimen, however, each site were able to make decisions regarding enrollment criteria. The Toronto location (hereafter referred to as IELCAP-Toronto), was based out of Princess Margaret Cancer Centre and began in 2003. IELCAP-Toronto enrolled 4,782 adults age 50 or older who were ever-smokers with more than 10 pack-years history of smoking. Participants were screened at baseline with milt-detector-row CT scanners. Positive findings were considered as any NCN found on a baseline scan.

### Pittsburgh Lung Screening Study (PLuSS)

PLuSS was a lung cancer screening trial that recruited 3,642 eligible participants between January 2002 and April 2005^30^. Eligible participants included those age 50 to 79 years, with no personal history of lung cancer, no concurrent participation in other lung screening studies, no chest CT within the preceding year, current or former smoker with 0.5 pack-years history of smoking for at least 25 years, no smoking cessation within 10 years of enrollment, and body weight less than 400 pounds. Participants underwent lose-dose chest CT at baseline. Positive findings were considered as any NCN.

### Pulmonary Nodule Segmentation

We performed supervised, semi-automated segmentation of screen-detected pulmonary nodules using the open-source 3D Slicer software^32^ and the Chest Imaging Platform extension^33, 34^. Our radiologist (HAS) located and reviewed each pulmonary lesion. Upon locating the lesion, the radiologist placed a seed-point at the approximate centroid of the lesion; semi-automated segmentation was performed based on the single seed-point, and manual touch-ups were performed at the discretion of the radiologist to fix over-or under-segmentation. All nodules were reviewed using standard lung windows. Segmentations for PanCan were performed by the PanCan investigators using an automated segmentation algorithm based on a commercial software and images and masks were provided without further processing, except those relevant to the feature extraction, detailed in the following section. We also collected detailed nodule information, including: lung and lobe location, suspicion of nodule malignancy, a nodule-specific LungRADS score (based on LungRADS 1.1,^8^), and ratings for semantic nodule features including: margin, sphericity, subtlety, spiculation, solidity, calcification, structure, and lobulation. Details on the ratings systems for semantic nodule features are described in **Supplementary Table 1**.

### Radiomics Feature Extraction

We performed radiomic feature extraction for baseline screen-detected pulmonary nodules using PyRadiomics (version 3.0.1)^26^. Due to heterogeneity in image acquisition settings between and within screening studies, all images and masks were resampled and interpolated to have unit voxel spacing (i.e., isotropy). We used a linear interpolator for images and nearest-neighbours interpolator for masks (to preserve labels). Grayscale intensities were discretized into bins using a bin width of 25 for histogram-based features. Voxel intensities were right-shifted by 1000 units prior to feature extraction to avoid negative values during feature computations.

Feature classes and the number of features per class were: (1) first-order statistics [18 features], (2) shape-based [14 features], (3) gray level coocurrence matrix [24 features], (4) gray level run length matrix [16 features], (5) gray level size zone matrix [16 features], (6) neighbouring gray tone difference matrix [5 features], and (7) gray level dependence matrix [14 features]. The list of radiomic features for each class is provided in **Supplemental Table 2**. We extracted shape and intensity-based features using the original image. We also extracted intensity-based features from images after applying several transformations, including: wavelet, Laplacian of Gaussian (LoG), Square, SquareRoot, Logarithm, Exponential, Gradient, and LocalBinaryPattern3D. In total, we extracted 2,060 radiomic features per nodule.

## Statistical Analysis

### Epidemiologic covariates and outcomes

Epidemiologic data were harmonized across the four screening studies to establish a common set of patient-level covariates. After harmonization, age, sex, family history of lung cancer among a first-degree relative, history of COPD or emphysema, smoking status, smoking duration, smoking intensity, years since quitting, and body mass index were included. We combined epidemiologic and radiomic features from the four screening studies to form our candidate predictor set. Nodule malignancy status was the outcome of interest and was determined based on the nodule-specific radiological assessment described in the Supplemental Methods.

### Model Development

We used subject-level random sampling to split the data into training (80%) and testing (20%) sets, ensuring all nodules for a specific participant were in the same split. The training set was further split into five folds using subject-level random sampling to perform cross-validation. We had 2,060 radiomic features to assess for their ability to classify benign and malignant pulmonary nodules. Many radiomic features have correspondences to established clinically-collected (i.e., semantic) nodules features, however, many features have an unknown predictive value. We performed an initial set of filtering steps to remove zero variance (n=78), low quality (n=11), and weakly predictive (FDR-adjusted P-value > 0.05 in univariate models, n=248), and highly-redundant features (pairwise correlation > 0.9, n=1,081), described in detail in the Supplemental Methods.

Using the 9 epidemiologic covariates and 647 radiomic features retained after filtering, we performed cross-validation to identify the top-performing ML model. All predictors were normalized prior to model fitting. We assessed the following ML models: penalized logistic regression (LASSO), Random Forest (RF), and Gradient Boosted Trees (XGBoost). We first performed grid-search over a set of hyperparameters chosen using a Latin hypercube space-filling design^35^. We then performed random grid search over a finer set of hyperparameters for the top-performing model. The optimal hyperparameter(s) were then fit to the full training set and model performance was evaluated in the hold-out test set. A schematic of the analytic approach used in this study is presented in **Figure 1**. All statistical analysis was performed using Python 3.7.10 and R 4.0.5^36, 37^.

**Figure 1.**
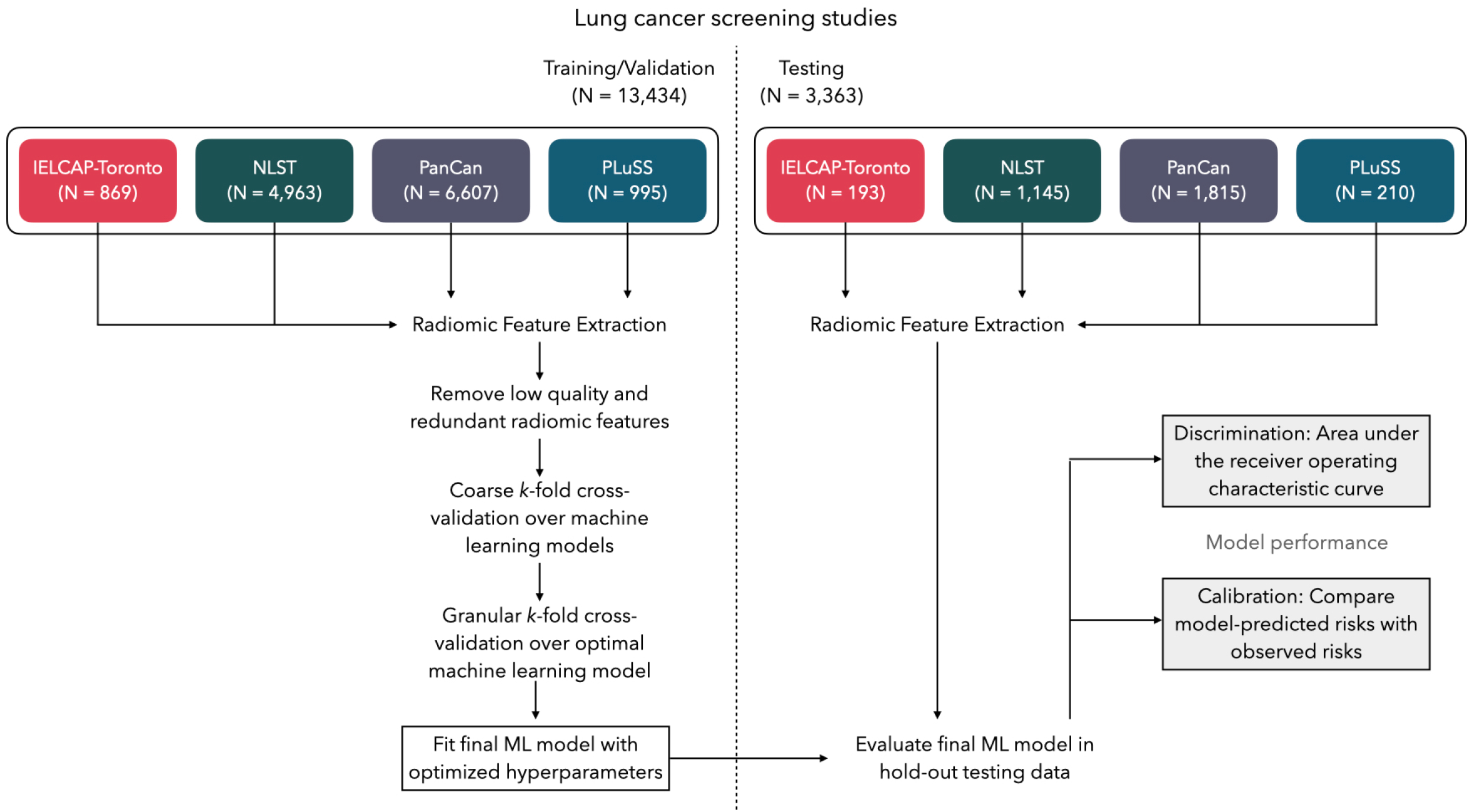
Schematic for the analytic framework used in this study. Data were partitioned into training/validation and testing splits using group-based random sampling to ensure all nodules for a participant were in a single set to avoid data leakage. Radiomic features were extracted and subject to filtering to exclude low-quality and highly-redundant features. K-fold cross-validation was performed to identify the optimal machine learning (ML) model and the optimal set of hyperparameters. The final ML model was fitted to the entire training data set and tested for out-of-sample performance in the hold-out test data; discrimination and calibration performance metrics are reported.

### Model Performance

We evaluated model performance in two complementary ways: (1) area under the receiver operating characteristic curve (AUC) to assess a models ability to assign higher risks to malignant lesions than to benign lesions (i.e., discrimination), and (2) compare model-estimated risks to observed risks (i.e., calibration). For calibration, we compared predicted and observed risks within quantiles of predicted risks, and also assessed the ratio of expected to observed number of cancers and the difference between expected and observed number of cancers. We report the AUC and calibration metrics with percentile-based bootstrap confidence intervals. We compared our model against an established nodule malignancy model (see Supplementary Materials for details).

## Results

Basic demographics about the participants and nodules in the four lung cancer screening cohorts are presented in **Table 1**. Participants were similar in age between the cohorts. There were more men than women in NLST (57% vs. 43%), PanCan (53% vs. 47%), and PLuSS (51% vs. 49%), while IELCAP-Toronto (61% vs. 39%) had more women. The four cohorts had differing proportions of current and former smokers, and smoking histories (i.e., duration, intensity, and years since quitting) varied between studies. All four cohorts generally consisted of heavy current and former smokers. On average, PanCan had more nodules per participant, and smaller nodules, compared to the other studies.

**Table 1.**
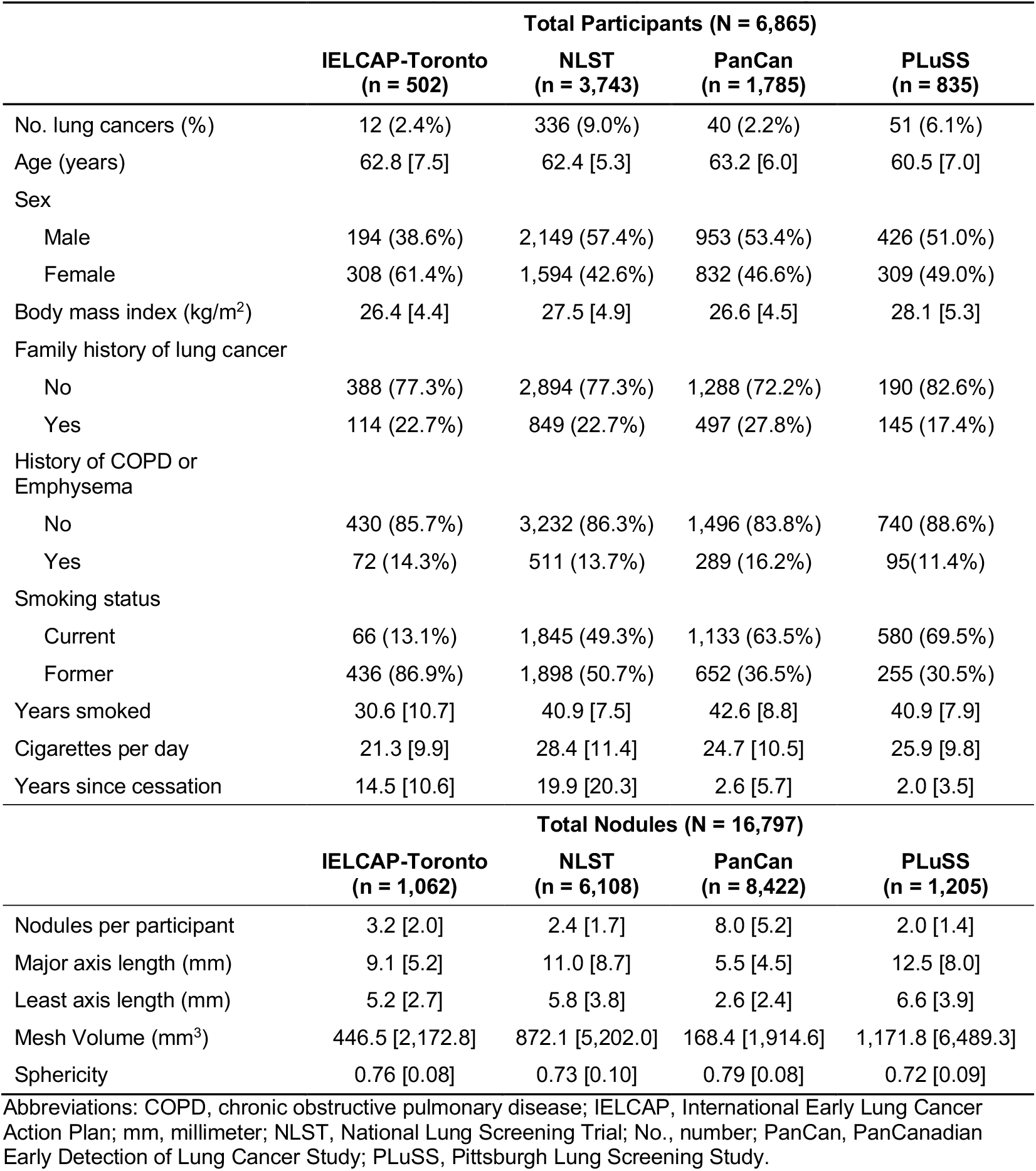
Patient-level and nodule-level descriptive statistics for each of the four screening cohorts included in this study. Means and standard deviations are reported for numeric variables and counts and proportions are reported for categorical variables.

We excluded 1,284 nodules from our study due technical issues with feature extraction, 2,574 nodules not first-appearing on baseline scans, and another 2,103 nodules due to missing patient-level data for the harmonized set of epidemiologic covariates. In total, we had 16,797 baseline screen-detected nodules among 6,865 participants for our analytic sample. A complete flow chart for nodule inclusion in the analytic sample is presented in **Supplemental Figure 1**. Distributional measures for the radiomic features based on the original CT image are presented in **Supplemental Table 4**.

We started with 2,060 radiomics features for model development. We removed 78 features due to zero-variance and 11 features due to observed numerical instability (i.e., implausible values) for a large number of participants. Next, we fit univariate models for each feature in the training data, and retained features with a FDR-adjusted p-value less than 0.05 (n=248). Lastly, we evaluated all pairwise sets of predictors with correlation in the training set greater than 0.9 (in descending order) and removed the predictor with the larger p-value. We retained 642 radiomic features for model development. More details can be found in the Supplementary Materials and **Supplemental Figure 2**. The 642 radiomics features retained for model development are presented in **Supplemental Table 4**. We performed unsupervised clustering in the training data set using the 642 radiological features which revealed three distinct clusters of participants with similar radiomics profiles (see **Supplemental Figure 3**). We compared the three clusters based on their proportions of malignant pulmonary nodules and found statistically significant differences (P_Exact_ < 0.05).

We fit three different machine learning models (LASSO, XGBoost, Random Forest) using 5-fold cross-validation based on the 642 radiomics features and 9 epidemiologic covariates. We first fit a coarse grid of 50 sets of hyperparameters for each ML model. The results for this first-pass cross-validation are presented in **Table 2** and **Supplemental Figure 4 and 5**. We selected the top performing model (LASSO) based on the combination of discrimination (AUC) and calibration (calibration ratio) and performed a final cross-validation and grid search over a finer grid of hyperparameters. The optimal penalty value for the LASSO, based on CV, was used to fit the final model based on the full training data set, and predictions were made on the held-out test set to evaluate model performance.

**Table 2.** Area under the receiver operating characteristic curve (AUC) based on the K-fold cross-validation of three different machine learning classification models for nodule malignancy prediction based on epidemiologic and radiomic features. We present the cross-validated AUC and confidence intervals.

| **ML Model** | **Optimal hyperparameters** | **CV-AUC (95% CI)** |
| --- | --- | --- |
| XGBoost | Num. of trees = 149<br>Tree depth = 11<br>Minimum node size = 15<br>Num. of predictors = 452<br>Learning rate = 0.0673<br>Loss reduction = 4.315 | 0.933 (0.923-0.944) |
| LASSO <sup>1</sup> | Penalty = 0.00044 | 0.930 (0.914-0.946) |
| Random Forest | Num. of trees = 147<br>Num. of predictors = 53<br>Minimum node size = 26 | 0.916 (0.904-0.929) |

Abbreviations: AUC, area under the curve; CI, confidence interval; LASSO, least absolute shrinkage and selection operator; ML, machine learning; Num, number; XGBoost, eXtreme Gradient Boosting.
<sup>1</sup> The penalty parameter for the LASSO model was a L1 (i.e., LASSO) penalty.

The top ML submodels that yielded the highest cross-validated AUC were XGBoost (AUC=0.933, 95% CI: 0.923-0.944), LASSO (AUC=0.930, 95% CI: 0.914-0.946), and Random Forest (AUC=0.916, 95% CI: 0.904-0.929). However, calibration was superior for the LASSO model and was chosen as the top model (see **Supplemental Figure 6**). In total, 142 predictors were retained in the final LASSO model with non-zero coefficients (See **Supplemental Figure 7**).

We compared our model with the established Brock Model. Our radiomics model had better discrimination, with a test-set AUC of 0.93 (95% CI: 0.90-0.96) compared to 0.87 (95% CI: 0.85-0.89) for the Brock Model (see **Figure 2**). Our model demonstrated excellent calibration when comparing observed risks with model-predicted (i.e., expected) risks, within quintiles of predicted risk. Our model had superior calibration compared to the Brock Model (see **Supplemental Table 5** and **Supplemental Figure 8**). We estimated the observed and expected number of malignant nodules (per 100,000) for the Brock model and our radiomics model. Our model had excellent calibration ratios (Exp / Obs) of 1.02 (95% CI: 0.89-1.18) and calibration differences of 69.39 (95% CI: -399.67, 507.94), versus 1.25 (95% CI: 1.15,1.36) and 1172.89 (95% CI: 774.17, 1583.65) for the Brock Model, respectively. We compare clinically-relevant metrics (e.g., sensitivity, specificity, etc.) between our model and the Brock model in **Table 3**. At nearly every probability threshold, our model has higher sensitivity, specificity, positive predictive value (PPV), negative predictive value (NPV), and accuracy, while identifying fewer lesions as positive (i.e., suspicious), when compared to the Brock model.

**Figure 2.**
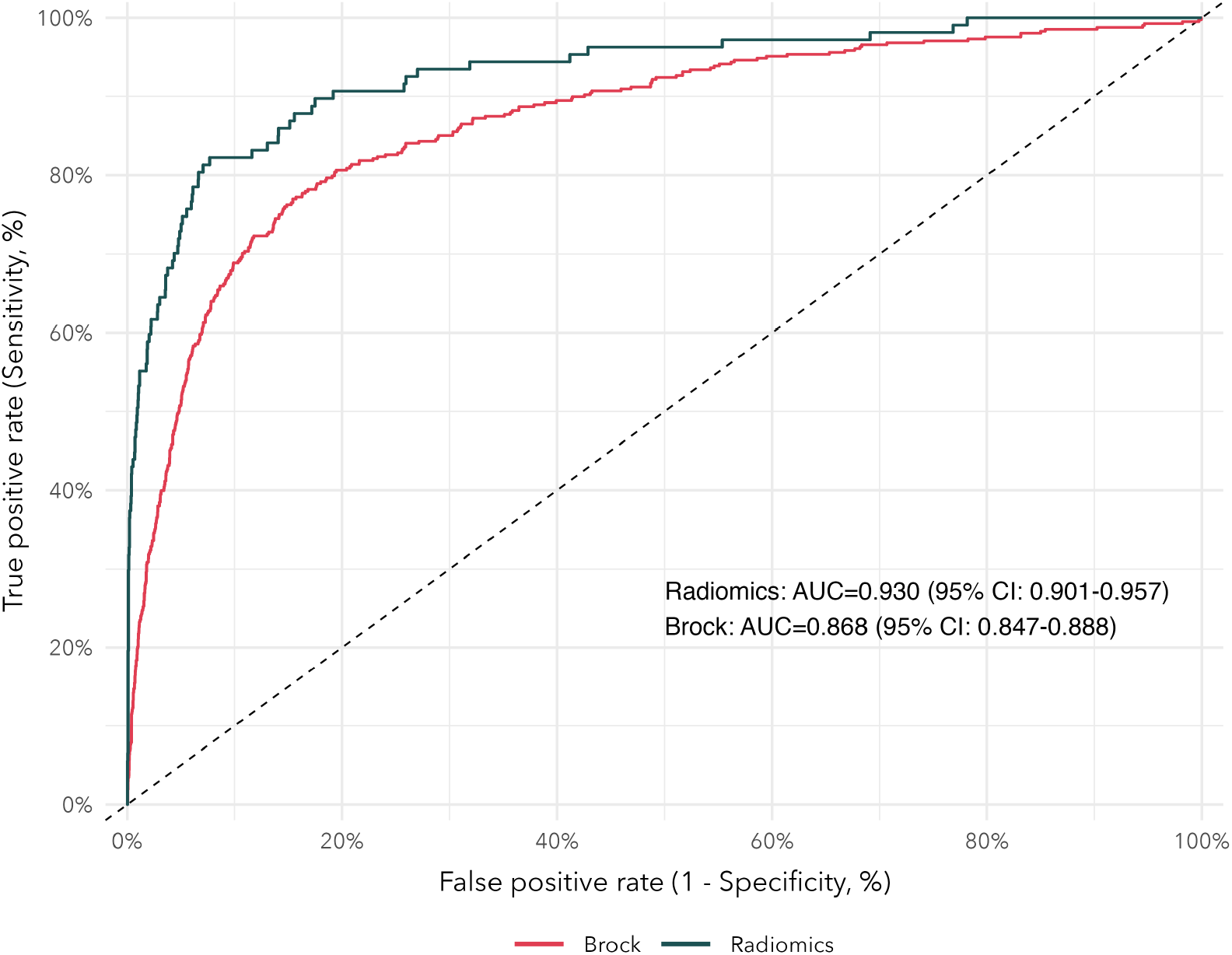
Receiver operating characteristic (ROC) curves for our radiomics models and the established Brock Model. Area under the curve (AUC) and 95% confidence intervals are reported.

**Table 3.** Comparison of sensitivity, specificity, positive predictive value, negative predictive value, accuracy, and positive prevalenc between our radiomics model and the established Brock Model. We provide point estimates and 95% percentile-based bootstrap confidence intervals for each statistic.

| Probability Threshold | Sensitivity (%) | Specificity (%) | PPV (%) | NPV (%) | Accuracy (%) | Positive Prevalence (%) |
| --- | --- | --- | --- | --- | --- | --- |
| Radiomics <sup>1</sup> |  |  |  |  |  |  |
| ≥2% | 89.7 (83.8-95.1) | 81.6 (80.3-82.9) | 13.8 (11.4-16.6) | 99.6 (99.4-99.8) | 81.9 (80.6-83.2) | 20.6 (19.4-22.0) |
| ≥5% | 82.2 (75.2-89.4) | 90.9 (89.9-91.8) | 23.0 (19.1-27.4) | 99.4 (99.1-99.6) | 90.7 (89.6-91.6) | 11.4 (10.4-12.5) |
| ≥10% | 74.8 (66.7-82.6) | 94.7 (94.0-95.5) | 31.9 (26.4-37.9) | 99.1 (98.8-99.5) | 94.1 (93.3-94.9) | 7.5 (6.6-8.4) |
| ≥15% | 64.5 (56.1-73.4) | 96.7 (96.1-97.3) | 39.2 (32.4-47.0) | 98.8 (98.4-99.2) | 95.7 (95.0-96.3) | 5.2 (4.5-6.0) |
| ≥20% | 61.7 (53.3-70.8) | 97.8 (97.2-98.3) | 47.5 (39.3-56.2) | 98.7 (98.4-99.1) | 96.6 (96.0-97.2) | 4.1 (3.4-4.8) |
| ≥25% | 55.1 (45.8-64.6) | 98.5 (98.0-98.9) | 54.6 (45.2-64.1) | 98.5 (98.1-98.9) | 97.1 (96.6-97.7) | 3.2 (2.6-3.8) |
| Brock Model <sup>2</sup> |  |  |  |  |  |  |
| ≥2% | 87.7 (84.7-90.6) | 64.5 (63.5-65.5) | 10.9 (9.9-12.1) | 99.1 (98.8-99.3) | 65.6 (64.7-66.6) | 38.0 (37.0-39.0) |
| ≥5% | 80.6 (76.9-84.0) | 79.8 (78.9-80.7) | 16.6 (15.0-18.3) | 98.8 (98.5-99.0) | 79.9 (79.0-80.7) | 23.0 (22.2-23.9) |
| ≥10% | 72.3 (68.1-76.5) | 88.0 (87.3-88.7) | 23.0 (20.6-25.6) | 98.5 (98.2-98.7) | 87.3 (86.5-88.0) | 14.9 (14.1-15.6) |
| ≥15% | 65.0 (60.6-69.4) | 91.6 (91.0-92.2) | 27.7 (24.9-30.7) | 98.1 (97.8-98.4) | 90.3 (89.7-91.0) | 11.1 (10.4-11.7) |
| ≥20% | 58.3 (53.6-63.0) | 93.7 (93.2-94.2) | 31.5 (28.2-34.9) | 97.8 (97.5-98.2) | 92.0 (91.5-92.6) | 8.8 (8.1-9.3) |
| ≥25% | 51.7 (47.0-56.3) | 94.9 (94.5-95.4) | 33.7 (30.0-37.4) | 97.5 (97.2-97.9) | 92.9 (92.4-93.4) | 7.3 (6.7-7.8) |

Abbreviations: NPV, negative predictive value; PPV, positive predictive value.
Note: Sensitivity is the proportion of malignant nodules correctly identified as malignant. Specificity is the proportion of benign nodules correctly identified as benign. PPV is the proportion of positive predictions that are malignant nodules. NPV is the proportion of negative predictions that are benign nodules. Accuracy is the total number of correct predictions out of the total number of nodules. Positive prevalence is the proportion of positive predictions divided by the total number of predictions.
<sup>1</sup> The radiomics model was evaluated in the 20% hold-out test data not used for model development (N = 3,363).
<sup>2</sup> The Brock Model was evaluated in the entire eligible set of participants from IELCAP-Toronto, NLST, and PLuSS (N = 8,622).

## Discussion

We developed and validated a pulmonary nodule malignancy assessment model based on radiomics and epidemiologic data from four large, international lung cancer screening cohorts using a machine learning approach. We found that the top-performing models were based on gradient boosted trees (XGBoost) and penalized logistic regression (LASSO), while the LASSO model provided the most optimal calibration. The use of quantitative imaging features (i.e., radiomics) showed improved performance compared to an established model based primarily on semantic nodule features. Radiomic features have demonstrated value for their ability to predict nodule malignancy risk and may improve the management of screen-detected pulmonary nodules by providing clinicians with supporting information for clinical decision-making.

Historically, the large quantity of medical images acquired during lung cancer screening have been under-utilized for extracting important information to inform nodule management. Traditionally, a modest set of semantic nodule traits are qualitatively assessed by expert radiologists to provide a high-level characterization of nodule morphology. High-throughput quantitative image analysis removes this layer of inter-reader subjectivity, while also collecting many more features that may further enhance our ability to characterize nodule morphology and intranodular textural heterogeneity^38^. Radiomic features can describe various aspects of the nodule morphology in ways that are imperceptible to the human eye (i.e., subtle intratumoral textural changes)^26^. The combination of radiomic features with known important patient-level features are expected to improve clinical management of nodules.

Previous studies demonstrated that quantitative image analysis can identify important prognostic signatures in head and neck cancer^38^. The feature extraction presented in^38^ was formalized as a free and open-source software^26^ and has enabled transparency and reproducibility for feature extraction, and contributed to the growing interest in quantitative image analysis in many areas of medical imaging, including lung cancer screening. To date, many of the radiomic studies for pulmonary nodule assessment have been performed based on relatively small data sets and with no ground-truth for nodule cancer status. Previous studies have shown that radiomic features can help identify lung cancer subtypes^39, 40^ and even the presence of therapy-targetable somatic mutations (e.g., EGFR, KRAS)^41–45^. The use of non-invasive image features is growing in popularity and will help improve lung cancer screening program efficiency.

To our knowledge, our radiomics study is the largest study to date to systematically investigate the importance of radiomics for pulmonary nodule assessment. We performed supervised, semi-automated segmentation of pulmonary nodules for three lung cancer screening studies using an open-source tool that is available for anyone to use. Our study was based on 16,797 nodules among 6,865 participants from four lung cancer screening cohorts. We used a systematic approach to develop machine learning prediction model using radiomics features that were consistently predictive across each of these four independent screening cohorts. With increasing usage of computer-aided diagnositc (CAD) software, the segmentation process can be fully automated. The model presented here can be easily implemented without additional processing need for a large-amount of images with the added advantage of minimum inter-reader variability.

Our study has several limitations worth highlighting. First, ground-truth nodule-level malignancy status was unavailable for two of the screening studies (NLST, PLuSS). As such, we used a set of rules to assign nodule-level malignancy status for participants with a lung cancer diagnosis. Imperfect assignment will lead to missclassification errors that can bias the results of our study. However, we used a relatively conservative approach based on suspicion of malignancy determined by expert review of nodules by our radiologist, who has extensive experience in lung CT assessment. For this reason, we believe the potential for missclassification bias is limited. There were feature extraction issues that excluded 5.7% of the candidate nodules. Nearly 80% of these issues were due to very small nodules with segmentation masks containing only a single voxel or were 1-dimensional after resampling and interpolation. These micronodules have a very low prior probability of being malignant and their exclusion are unlikely to bias our results. Lastly, there was numerical instability for a small set of radiomic features when computing on derived images (i.e., after transformations). We minimized potential bias from these unstable features by excluding them for the filters where identifiable problems arose. All radiomic features appeared stable based on the original image.

In summary, we developed a nodule assessment model based on quantitative imaging and patient-level features collected from four international lung cancer screening cohorts. We believe this study contributes important insights into the role that high-dimensional radiomic features can play in accurately assessing nodule malignancy risk and that these features generalize well to geo-temporally distinct screening cohorts. At present, there is emerging interest in analyzing medical images using deep learning computer vision approaches, although limited transparency in model development and lack of model interpretability can pose challenges for clinical implementation and widespread adoption^46, 47^. In the future, our model may help to improve nodule malignancy assessment and provide supplemental information that can help guide decision-making for screen-detected nodule management.

## Data Availability

All data used in the present study may be made available upon reasonable request to the Integrative Analysis of Lung Cancer Etiology and Risk (INTEGRAL) program upon approval by the Committee.

https://www.integralu19.org/proposals

## Notes

### Competing Interest Statement

The authors have declared no competing interest.

### Funding Statement

This work was supported by Canadian Institutes of Health Research (FDN 167273) and the National Institutes of Health (U19 CA203654).

### Author Declarations

Research ethics approval for all of the lung cancer screening studies included in the current study are covered by Mount Sinai Hospital (MSH) Research Ethics Board (REB) approval for the Integrative Analysis of Lung Cancer (INTEGRAL) project (MSH REB 17-0119-E).

