## Supplementary Materials for "Radiomics analysis to predict pulmonary nodule malignancy using machine learning approaches"

#### Supplementary Methods

**1 Computed tomography baseline screening protocol** **2**
1.1 National Lung Screening Trial 2
1.2 PanCanadian Early Detection of Lung Cancer Study 2
1.3 International Early Lung Cancer Action Program - Toronto 2
1.4 Pittsburgh Lung Screening Study 2

**2 Semi-automated pulmonary nodule segmentation** **3**

**3 Ground-truth nodule malignancy status** **5**

**4 Initial filtering of radiomics features** **6**

**5 Application of the Brock Model** **7**

**References** **8**

### 1 Computed tomography baseline screening protocol

The following sections will briefly describe the CT screening protocol used in each of the four lung cancer screening studies.

#### 1.1 National Lung Screening Trial

CT screening was done using multi-detector scanners (at least four detectors) for a helical acquisition with the participant in the supine position (arms elevated above the head) with suspended maximal inspiration with the following acquisition settings: 120 to 140 kVp voltage, 40 to 80 mA current, 2.5mm detector collimation, 1.0 to 3.2 mm nominal reconstructed section width, 1.0 to 2.5 reconstruction interval, 25 second scanning time, and a soft tissue or thin section reconstruction kernel. All low-dose CT scanners were certified for use in the NLST by meeting the acquisition requirements described previously. More details can be found in[1].

#### 1.2 PanCanadian Early Detection of Lung Cancer Study

Screening was performed during a single inspiratory breath hold with the participant in a supine position. PanCan used multi-detector row CT scanners (minimum of four detectors) with a reconstruction interval (i.e., slice thickness) in the trans-axial plane of 1.25mm or less, and within-plane spacing of 1.25mm or less. PanCan used different reconstruction algorithms (kernels), one for lung parenchyma, and another for visualizing mediastinal structures. Scans were performed with 120kV, 40 to 50 mAs, and a rotation time of less than 1 second. Each scan was reviewed by a designated radiologist for each participating centre. More details can be found in[2] and[3].

#### 1.3 International Early Lung Cancer Action Program - Toronto

Baseline CT scans for IELCAP-Toronto were acquired using multi-detector-row CT scanners with 16 or more rows and 4 to 64 channels. Low-dose, thin-slice CT scans were performed using the following acquisition settings: axial reconstruction slice thickness at 1.25mm or less, 120 kVp or lower, 40 to 60 mAs, and a non-edge enhancing image reconstruction kernel. Radiologists reviewed scans at the Toronto site, aware of which screening round (baseline or repeat) the scan belongs to. The primary objective for the baseline scans was to identify all non-calcified nodules. More details can be found in[4].

#### 1.4 Pittsburgh Lung Screening Study

Screening was performed using a single-breath-hold, helical, low-dose technique (40-60 mA, 140 kVp) to obtain axial reconstructions with slice thickness of 2.5mm intervals. PLuSS used a high-spatial frequency lung kernel at contiguous 2.5mm intervals. All scans were reviewed by one of two study radiologists using standard lung windows. More details can be found in[5].

#### 2 Semi-automated pulmonary nodule segmentation

We performed supervised semi-automated segmentations of screen-detected pulmonary nodules using the open-source 3D Slicer software[6] and the Chest Imaging Platform (CIP) extension[7]. The Lung Lesion Analyzer (LLA) module within CIP was used for nodule segmentations. The semi-automated segmentation algorithm has been previously validated against consensus segmentations performed manually by several trained radiologists and was found to perform similarly well[8].

Technical details about the segmentation algorithm used in the LLA are described in[9] and[8]. Here we provide a brief description of the algorithm. The segmentation algorithm used by the LLA is a level set algorithm[9]. The level set algorithm iteratively grows outward the boundary between the nodule and non-nodule, with the growth distance being proportional to the distance from the current front to a feature boundary[9]. These feature boundaries represent areas that the region growing algorithm should avoid. By default, the four feature boundaries are: (1) the lung wall, (2) the lung vasculature, (3) the boundary between the nodule and the background lung parenchyma, and (4) the low CT density of air. These boundaries allow the outward growth of the nodule segmentation, while deterring over-segmentation into the lung wall, vessel, or across sharp edges. In other words, the nodule segmentation iteratively grows, and the rate of growth slows down as the segmentation approaches parts of the lung that do not belong to the nodule. The boundary constraints improve upon other common segmentation algorithms, such as GrowCut, which can segment large tumor volumes with relative ease, but are less robust for smaller pulmonary lesions, such as those typically identified during lung cancer screening, or lesions with pleural and/or vessel attachments[9].

1. **Lung wall** - The lung wall feature is used to deter the nodule contour from growing beyond the lung wall and into the pleura, which is especially important when segmenting juxtapleural lesions. The lung wall can be characterized by a much higher voxel intensity than the lung parenchyma (which is primarily air). However, juxtapleural lesions may have voxel intensities quite similar to the lung wall. These juxtapleural lesions create interruptions in the otherwise smooth, consistent curvature of the lung wall. Thus, the lung wall feature is created by first binarizing the image at a threshold of -400 HU, and then, for voxels near the lung boundary, apply a hole-filling algorithm at a per-voxel level to decide whether a given voxel will be filled based on a majority-voting scheme using a neighborhood of 7x7x7 surrounding the candidate voxel. The hole-filling algorithm is ran over many iterations until no voxels labels get changed from background to foreground. This has the effect of reducing surfaces to the point of having low Gaussian curvature. Lastly, a sigmoid function is applied to the final binary image to map the values to a smooth and continuous range.

2. **Vesselness** - Pulmonary nodules are compact in size, while lung vasculature and vessels are well-approximated by a cylinder or tube. This feature computes how closely a given image region resembles a tube. This is done using the Sato tubularness measure. A sigmoid function is applied to the final binary image.

3. **Gradient** - The gradient feature is used to help identify the edges of the lesion when the lesion is surrounded by a lower density structure/area, such as the lung parenchyma. The well-established Canny edge detector is used to produce an edge map. A sigmoid function is applied to the final binary image.
4. **Intensity** - The intensity feature is based on a simple intensity threshold. Based on whether the nodule is solid or part-solid, a threshold of -200 HU or -500 HU is used for thresholding, respectively. A sigmoid transformation is applied after thresholding.

All of the features are aggregated after normalizing each feature to have values in the range of [0,1] and the aggregated image is used for segmentation. The segmentation algorithm is initialized by the user placing a single seed point inside the nodule volume, typically at the 3D centroid of the nodule. The nodule segmentation is then performed using the above described level set algorithm, based on a front propagation approach[9]. The propagation of the segmentation is constrained to prevent leakage of the segmentation into non-nodule volumes including the chest wall, airway walls, or other regions with tubular or vessel-like structures[9]. After the segmentation algorithm is complete, the user may perform manual touch-ups to fix any over- or under-segmentations.

### 3 Ground-truth nodule malignancy status

Both the PanCan and IELCAP-Toronto tracked malignancy status (i.e. lung cancer) at a per-nodule level. However, for PLuSS and NLST, lung cancer status was only known at the patient level. Our radiomics feature extraction and analysis is at the nodule level, so we employed a set of heuristics to prepare these cohorts for inclusion in this study. For NLST and PLUSS, among participants with lung cancer and annotated nodules, we selected the nodule with the highest suspicion of malignancy for inclusion. Suspicion was determined (in order) by: (1) radiologist assigned LungRADS 1.1 score, (2) malignancy rating, (3) longest axis, and (4) longest perpendicular axis. We only included participants whose lung cancer diagnosis date was within 2 years of their baseline screen to avoid including lesions with low probability of being the lesion that led to a lung cancer diagnosis.

### 4 Initial filtering of radiomics features

We performed a first-pass filter of the radiomics features to reduce the set of predictors for inclusion in the cross-validation and machine learning model development. Next, we describe the reasoning and application of each of the filtering steps for preliminary feature selection. The filtering steps were applied in the order as shown.

1. **Computational Issues**
    - First, we removed any predictors with zero-variance. In other words, predictors containing only a single unique value were removed as they would, by definition, have no predictive value in models due to lack of variability. Next, several variables were observed to be numerically unstable after applying some of the image filters (i.e., during feature extraction based on derived images). In particular, we removed the Coarseness (NGTDM) feature for all but the original and eight wavelet filters.
2. **Weak Predictors**
    - We fit univariate logistic regression models for each of the radiomic features that passed Step 1, with nodule malignancy as the response variable. We removed putatively weak predictors based on the p-value associated with the beta coefficient (i.e., the logit) from the logistic regression model. We adjusted the p-values using the Benjamini-Hochberg correction to control the false-discovery rate (FDR)[10]. Any radiomic feature with a univariate FDR-adjusted p-value of less than 0.05 were retained.
3. **Redundancy**
    - Lastly, we aimed to remove some of the redundancy among features due to extreme correlation. Several radiomic features are perfect correlates of one another and thus carry identical information for prediction purposes. We examined all pairwise sets of features with an absolute Pearson's correlation coefficient more than 0.9. We sorted the sets of predictors in descending order. Next, starting with the most highly correlated set of features, we removed one of the two features based on which features had the larger FDR-adjusted P-value. We iteratively performed this feature selection process until we had a pruned set of features with correlation of less than or equal to 0.9 (based on the absolute correlation).

There were also 1,284 pulmonary nodules excluded from our analytic sample due to technical issues with the feature extraction. Technical issues included: (1) missing or corrupt mask files (n=3), (2) issues with computing shape statistics (n=88), (3) geometric mismatches (n=93), (4) masks contained only a single voxel (n=448), (5) masks were 1-dimensional (n=540), or (6) removed nodules with sentinel values for incalculable features (n=112).

#### 5 Application of the Brock Model

The Brock model is an established nodule malignancy model developed based on 1,871 participants from the PanCanadian Early Detection of Lung Cancer Study (PanCan)[2]. The original paper describes four models, named as Model 1a (Parsimonious, No Spiculation), Model 2a (Full Model, No Spiculation), Model 1b (Parsimonious, with Spiculation), and Model 2b (Full Model, with Spiculation). The parsimonious models included sex (female vs. male), nodule size (mm), and nodule location (upper vs. middle or lower lobe). The full model further included age (years), family history of lung cancer (yes vs. no), emphysema (yes vs. no), nodule size (mm), nodule type (Nonsolid or with ground-glass opacity, Part-solid, Solid), and nodule count per scan (per additional nodule). Models 1a and 2a did not included spiculation (yes vs. no), while Models 1b and 2b included this variable. Nodule size is included in the models as a non-linear transformation based on multiple fractional polynomials. The functional form of nodule size (per mm) is as follows: $\left(\left(\frac{\texttt{Nodule size}}{10}\right)^{-0.5}\right) - 1.58113883$. Age was centered at 62 years, nodule size is centered at 4mm, and nodule count per scan was centered at 4 nodules.

We applied Model 2b (Full Model, with Spiculation) in our data set as this is the fullest model containing all of the predictors. Since the Brock Model was developed in PanCan, we only apply the Brock Model to the NLST, IELCAP-Toronto, and PLuSS to avoid presenting overly-optimistic performance metrics. The risk formula for the Brock Models is:

$$\begin{aligned}\texttt{Brock Logit} = {} & -6.7892 + ((\texttt{Age} - 62) * 0.0287) + (\texttt{Female} * 0.6011) + (\texttt{FHLC} * 0.2961) + \\ & (\texttt{COPD} * 0.2953) + \left(\left(\left((\frac{\texttt{Nodule size}}{10})^{-0.5}\right) - 1.58113883\right) * -5.3854\right) + \\ & (\texttt{GGO} * -0.1276) + (\texttt{Part solid} * 0.3770) + (\texttt{Upper lobe} * 0.6581) + \\ & ((\texttt{Nodule count} - 4) * -0.0824) + (\texttt{Spiculation} * 0.7729)\end{aligned}$$

Where Female, FHLC, COPD, GGO, Part Solid, Upper Lobe, and Spiculation are binary indicator variables for the presence (1) or absence (0) of the trait. We can then get the probability as:

$$\texttt{Brock Prob} = \frac{\exp(\texttt{Brock Logit})}{(1 + \texttt{Brock Logit})}$$

**Supplemental Table 1.** Ratings systems used for collecting semantic nodule features for IELCAP-Toronto, PanCan, and PLuSS.

| **Feature** | **Rating System** | | | | | |
|---|---|---|---|---|---|---|
| Calcification | 1=popcorn | 2=laminated | 3=solid | 4=non-central | 5=central | 6=absent |
| Internal structure | Soft tissue | Fluid | Fat | Air | | |
| Lobulation | 1=none | 2 | 3 | 4 | 5=marked | |
| Malignancy | 1=highly unlikely | 2=moderately unlikely | 3=indeterminate | 4=moderately suspicious | 5=highly suspicious | |
| Margin | 1=poorly defined | 2 | 3 | 4 | 5=sharp | |
| Radiographic solidity | 1=non-solid/GGO | 2 | 3=part-solid/mixed | 4 | 5=solid | |
| Sphericity | 1=linear | 2 | 3=ovoid | 4 | 5=round | |
| Subtlety | 1=extremely subtle | 2=moderately subtle | 3=fairly subtle | 4=moderately obvious | 5=obvious | |
| Spiculation | 1=none | 2 | 3 | 4 | 5=marked | |

Note: The rating system was based on the semantic features described in The Lung Image Database Consortium (LIDC) Data Collection Process for Nodule Detection and Annotation; McNitt-Gray MF, Armato III SG, Meyer CR, Reeves AP, McLennan G, Pais RC, Freymann J, Brown MS, Engelmann RM, Bland PH, Laderach GE. The Lung Image Database Consortium (LIDC) data collection process for nodule detection and annotation. Academic radiology. 2007 Dec 1;14(12):1464-74.

**Supplemental Table 2.** Radiomic features extracted based on the PyRadiomics library.

| First-order statistics | Shaped-based (3D) | Gray level cooccurrence matrix | Gray level size zone matrix | Gray level run length matrix | Neighbouring gray tone difference matrix | Gray level dependence matrix |
|---|---|---|---|---|---|---|
| Energy | Mesh volume | Autocorrelation | Small Area Emphasis (SAE) | Short Run Emphasis (SRE) | Coarseness | Small Dependence Emphasis (SDE) |
| Total Energy | Voxel volume | Joint Average | Large Area Emphasis (LAE) | Long Run Emphasis (LRE) | Contrast | Large Dependence Emphasis (LDE) |
| Entropy | Surface Area | Cluster Prominence | Gray Level Non-Uniformity (GLN) | Gray Level Non-Uniformity (GLN) | Busyness | Gray Level Non-Uniformity (GLN) |
| Minimum | Surface area to volume ratio | Cluster Shade | Gray Level Non-Uniformity Normalized (GLNN) | Gray Level Non-Uniformity Normalized (GLNN) | Complexity | Dependence Non-Uniformity (DN) |
| 10$^{th}$ percentile | Sphericity | Cluster Tendency | Size-Zone Non-Uniformity (SZN) | Run Length Non-Uniformity (RLN) | Strength | Dependence Non-Uniformity Normalized (DNN) |
| 90$^{th}$ percentile | Maximum 3D diameter | Contrast | Size-Zone Non-Uniformity Normalized (SZNN) | Run Length Non-Uniformity Normalized (RLNN) | | Gray Level Variance (GLV) |
| Maximum | Maximum 2D diameter (Slice) | Correlation | Zone Percentage (ZP) | Run Percentage (RP) | | Dependence Variance (DV) |
| Mean | Maximum 2D diameter (Column) | Difference Average | Gray Level Variance (GLV) | Gray Level Variance (GLV) | | Dependence Entropy (DE) |
| Median | Maximum 2D diameter (Row) | Difference Entropy | Zone Variance (ZV) | Run Variance (RV) | | Low Gray Level Emphasis (LGLE) |
| Interquartile range | Major axis length | Difference Variance | Zone Entropy (ZE) | Run Entropy (RE) | | High Gray Level Emphasis (HGLE) |
| Range | Minor axis length | Joint Energy | Low Gray Level Zone Emphasis (LGLZE) | Low Gray Level Run Emphasis (LGLRE) | | Small Dependence Low Gray Level Emphasis (SDLGLE) |
| Mean absolute deviation (MAD) | Least axis length | Joint Entropy | High Gray Level Zone Emphasis (HGLZE) | High Gray Level Run Emphasis (HGLRE) | | Small Dependence High Gray Level Emphasis (SDHGLE) |
| Robust mean absolute deviation (rMAD) | Elongation | Informational Measure of Correlation (IMC) 1 | Small Area Low Gray Level Emphases (SALGLE) | Small Run Low Gray Level Emphases (SRLGLE) | | Large Dependence Low Gray Level Emphasis (LDLGLE) |
| Root mean squared (RMS) | Flatness | Informational Measure of Correlation (IMC) 2 | Small Area High Gray Level Emphasis (SAHGLE) | Small Run High Gray Level Emphasis (SRHGLE) | | Large Dependence High Gray Level Emphasis (LDHGLE) |
| Skewness | | Inverse Difference Moment (IDM) | Large Area Low Gray Level Emphasis (LALGLE) | Large Run Low Gray Level Emphasis (LRLGLE) | | |

| | | | |
|---|---|---|---|
| Kurtosis | Maximal Correlation Coefficient (MCC) | Large Area High Gray Level Emphasis (LAHGLE) | Large Run High Gray Level Emphasis (LRHGLE) |
| Variance | Inverse Difference Moment Normalized (IDMN) | | |
| Uniformity | Inverse Difference (ID) | | |
| | Inverse Difference Normalized (IDN) | | |
| | Inverse Variance | | |
| | Maximum Probability | | |
| | Sum Average | | |
| | Sum Entropy | | |
| | Sum of Squares | | |

**Supplemental Table 3.** Descriptive statistics for the distribution of radiomic features across all four lung cancer screening studies.

| Feature | Mean | SD | Q25 | Median | Q75 | Min | Max |
|---|---|---|---|---|---|---|---|
| **First Order** | | | | | | | |
| 10Percentile | -578.30 | 189.20 | -737.10 | -599.84 | -435.40 | -1783.00 | 813.90 |
| 90Percentile | -146.79 | 327.52 | -348.70 | -141.20 | 14.00 | -1036.30 | 2235.00 |
| Energy | 342222389.86 | 3569036686.67 | 7171515.00 | 24780211.00 | 77743212.00 | 182154.00 | 249241111529.92 |
| Entropy | 3.90 | 1.02 | 3.21 | 4.12 | 4.70 | 0.57 | 6.87 |
| InterquartileRange | 247.27 | 157.24 | 137.95 | 225.50 | 321.00 | 0.00 | 1814.25 |
| Kurtosis | 2.52 | 1.26 | 2.01 | 2.28 | 2.69 | 1.00 | 50.66 |
| Maximum | -5.73 | 404.09 | -232.00 | -17.00 | 127.60 | -841.62 | 3006.00 |
| Mean | -366.85 | 233.17 | -537.80 | -364.72 | -224.57 | -1468.31 | 1165.78 |
| MeanAbsoluteDeviation | 141.91 | 84.85 | 85.88 | 131.07 | 178.43 | 6.15 | 907.36 |
| Median | -372.63 | 237.70 | -558.00 | -372.00 | -215.50 | -1515.50 | 1423.00 |
| Minimum | -698.77 | 200.11 | -852.00 | -739.00 | -549.00 | -3024.00 | 783.00 |
| Range | 693.05 | 422.36 | 400.00 | 646.00 | 889.00 | 23.00 | 3843.00 |
| RobustMeanAbsoluteDeviation | 101.34 | 65.66 | 57.38 | 93.65 | 132.36 | 0.00 | 756.97 |
| RootMeanSquared | 659.74 | 243.37 | 490.53 | 662.72 | 798.61 | 97.73 | 2364.31 |
| Skewness | 0.11 | 0.57 | -0.21 | 0.13 | 0.45 | -6.04 | 4.32 |
| TotalEnergy | 342237726.54 | 3569035354.26 | 7190243.00 | 24798216.00 | 77772859.00 | 182154.00 | 249241111529.92 |
| Uniformity | 0.10 | 0.08 | 0.04 | 0.07 | 0.12 | 0.01 | 0.81 |
| Variance | 39243.61 | 64258.38 | 10959.85 | 25036.20 | 45340.70 | 62.85 | 1017823.06 |
| **Gray Level Co-occurrence Matrix (GLCM)** | | | | | | | |
| Autocorrelation | 331.99 | 420.22 | 78.36 | 215.07 | 443.75 | 1.17 | 12595.05 |
| ClusterProminence | 189793.50 | 1208786.45 | 2302.68 | 19723.28 | 82202.64 | 0.00 | 40054589.22 |
| ClusterShade | 133.85 | 4914.62 | -266.72 | -3.67 | 69.64 | -202132.69 | 138982.72 |
| ClusterTendency | 141.10 | 236.69 | 28.09 | 84.36 | 178.03 | 0.00 | 3926.45 |
| Contrast | 98.68 | 212.14 | 28.18 | 53.48 | 92.44 | 0.17 | 3891.90 |
| Correlation | 0.11 | 0.42 | -0.14 | 0.19 | 0.43 | -1.00 | 0.94 |
| DifferenceAverage | 6.62 | 4.48 | 4.22 | 5.84 | 7.70 | 0.17 | 49.15 |
| DifferenceEntropy | 2.85 | 1.28 | 2.04 | 3.27 | 3.80 | 0.00 | 5.97 |
| DifferenceVariance | 28.88 | 64.37 | 6.85 | 16.09 | 28.78 | 0.00 | 1262.46 |
| Id | 0.26 | 0.09 | 0.21 | 0.25 | 0.30 | 0.03 | 0.92 |
| Idm | 0.18 | 0.10 | 0.12 | 0.16 | 0.21 | 0.00 | 0.92 |
| Idmn | 0.92 | 0.06 | 0.89 | 0.93 | 0.95 | 0.60 | 1.00 |
| Idn | 0.82 | 0.06 | 0.78 | 0.83 | 0.86 | 0.55 | 0.98 |
| Imc1 | -0.57 | 0.26 | -0.80 | -0.57 | -0.36 | -1.00 | -0.01 |
| Imc2 | 0.95 | 0.07 | 0.94 | 0.98 | 1.00 | 0.15 | 1.00 |
| InverseVariance | 0.18 | 0.10 | 0.12 | 0.16 | 0.22 | 0.00 | 1.00 |
| JointAverage | 15.56 | 8.80 | 8.89 | 14.39 | 20.54 | 1.08 | 111.61 |
| JointEnergy | 0.09 | 0.15 | 0.01 | 0.02 | 0.10 | 0.00 | 0.80 |
| JointEntropy | 5.38 | 2.39 | 3.44 | 5.81 | 7.28 | 0.40 | 12.22 |
| MaximumProbability | 0.84 | 0.15 | 0.72 | 0.88 | 1.00 | 0.10 | 1.00 |

| | | | | | | | |
|---|---|---|---|---|---|---|---|
| MCC | 0.11 | 0.14 | 0.02 | 0.04 | 0.13 | 0.00 | 0.83 |
| SumAverage | 31.12 | 17.60 | 17.78 | 28.78 | 41.08 | 2.17 | 223.23 |
| SumEntropy | 3.66 | 1.79 | 2.33 | 4.18 | 5.12 | 0.00 | 7.81 |
| SumSquares | 59.94 | 105.08 | 16.18 | 37.38 | 67.72 | 0.07 | 1549.00 |
| **Gray Level Dependence Matrix (GLDM)** | | | | | | | |
| DependenceEntropy | 4.74 | 1.51 | 3.62 | 5.01 | 5.91 | 0.81 | 8.31 |
| DependenceNonUniformity | 103.57 | 555.67 | 9.33 | 26.40 | 65.60 | 1.67 | 35608.44 |
| DependenceNonUniformityNormalized | 0.44 | 0.22 | 0.30 | 0.38 | 0.52 | 0.04 | 1.00 |
| DependenceVariance | 1.48 | 3.16 | 0.27 | 0.72 | 1.35 | 0.00 | 61.12 |
| GrayLevelNonUniformity | 41.78 | 728.25 | 1.91 | 4.29 | 9.58 | 1.00 | 69111.30 |
| GrayLevelVariance | 62.86 | 102.79 | 17.60 | 40.13 | 72.63 | 0.10 | 1628.29 |
| HighGrayLevelEmphasis | 330.94 | 416.98 | 87.78 | 213.50 | 427.63 | 1.43 | 10962.68 |
| LargeDependenceEmphasis | 5.95 | 9.08 | 2.43 | 3.81 | 5.88 | 1.00 | 227.44 |
| LargeDependenceHighGrayLevelEmphasis | 2919.93 | 10415.08 | 321.75 | 890.24 | 2011.22 | 3.00 | 364026.71 |
| LargeDependenceLowGrayLevelEmphasis | 0.25 | 0.91 | 0.04 | 0.09 | 0.21 | 0.00 | 50.31 |
| LowGrayLevelEmphasis | 0.08 | 0.10 | 0.02 | 0.04 | 0.10 | 0.00 | 0.89 |
| SmallDependenceEmphasis | 0.59 | 0.20 | 0.47 | 0.58 | 0.71 | 0.02 | 1.00 |
| SmallDependenceHighGrayLevelEmphasis | 189.25 | 318.54 | 46.99 | 114.11 | 215.81 | 0.11 | 7085.44 |
| SmallDependenceLowGrayLevelEmphasis | 0.06 | 0.08 | 0.01 | 0.03 | 0.08 | 0.00 | 0.57 |
| **Gray Level Run Length Matrix (GLRLM)** | | | | | | | |
| GrayLevelNonUniformity | 28.92 | 375.87 | 1.85 | 4.13 | 9.08 | 1.00 | 33004.92 |
| GrayLevelNonUniformityNormalized | 0.10 | 0.08 | 0.04 | 0.07 | 0.12 | 0.01 | 0.74 |
| GrayLevelVariance | 62.99 | 102.84 | 17.76 | 40.39 | 72.58 | 0.15 | 1628.00 |
| HighGrayLevelRunEmphasis | 327.99 | 414.23 | 87.80 | 212.85 | 423.01 | 1.54 | 10954.98 |
| LongRunEmphasis | 1.15 | 1.15 | 1.06 | 1.10 | 1.15 | 1.00 | 147.39 |
| LongRunHighGrayLevelEmphasis | 386.00 | 512.17 | 98.66 | 235.67 | 480.35 | 2.36 | 12031.22 |
| LongRunLowGrayLevelEmphasis | 0.09 | 0.14 | 0.02 | 0.04 | 0.11 | 0.00 | 10.00 |
| LowGrayLevelRunEmphasis | 0.08 | 0.10 | 0.02 | 0.04 | 0.10 | 0.00 | 0.86 |
| RunEntropy | 4.04 | 1.06 | 3.34 | 4.26 | 4.87 | 0.92 | 7.02 |
| RunLengthNonUniformity | 381.19 | 2310.97 | 17.33 | 65.28 | 177.17 | 2.00 | 124834.97 |
| RunLengthNonUniformityNormalized | 0.93 | 0.05 | 0.91 | 0.94 | 0.96 | 0.42 | 1.00 |
| RunPercentage | 0.96 | 0.03 | 0.95 | 0.97 | 0.98 | 0.52 | 1.00 |
| RunVariance | 0.05 | 0.39 | 0.02 | 0.03 | 0.05 | 0.00 | 49.00 |
| ShortRunEmphasis | 0.97 | 0.03 | 0.96 | 0.98 | 0.99 | 0.65 | 1.00 |
| ShortRunHighGrayLevelEmphasis | 317.03 | 404.33 | 85.34 | 207.11 | 409.83 | 1.34 | 10694.45 |
| ShortRunLowGrayLevelEmphasis | 0.08 | 0.09 | 0.02 | 0.04 | 0.10 | 0.00 | 0.66 |
| **Gray Level Size Zone Matrix (GLSZM)** | | | | | | | |
| GrayLevelNonUniformity | 7.63 | 32.08 | 1.33 | 2.62 | 5.10 | 1.00 | 1603.45 |
| GrayLevelNonUniformityNormalized | 0.09 | 0.08 | 0.04 | 0.06 | 0.11 | 0.01 | 0.56 |
| GrayLevelVariance | 64.02 | 103.39 | 19.91 | 42.00 | 71.52 | 0.22 | 1654.18 |
| HighGrayLevelZoneEmphasis | 302.91 | 394.91 | 87.50 | 202.30 | 380.20 | 2.00 | 10976.70 |
| LargeAreaEmphasis | 242.78 | 10666.83 | 1.88 | 3.00 | 5.44 | 1.00 | 1226672.91 |
| LargeAreaHighGrayLevelEmphasis | 348968.57 | 17648745.70 | 273.00 | 763.88 | 1870.58 | 3.00 | 2067360221.79 |

| | | | | | | | |
|---|---|---|---|---|---|---|---|
| LargeAreaLowGrayLevelEmphasis | 1.48 | 63.16 | 0.05 | 0.10 | 0.24 | 0.00 | 7771.40 |
| LowGrayLevelZoneEmphasis | 0.09 | 0.10 | 0.02 | 0.05 | 0.12 | 0.00 | 0.75 |
| SizeZoneNonUniformity | 96.54 | 507.46 | 8.20 | 23.94 | 60.58 | 1.00 | 30043.86 |
| SizeZoneNonUniformityNormalized | 0.58 | 0.18 | 0.47 | 0.56 | 0.68 | 0.06 | 1.00 |
| SmallAreaEmphasis | 0.77 | 0.13 | 0.70 | 0.77 | 0.84 | 0.02 | 1.00 |
| SmallAreaHighGrayLevelEmphasis | 235.76 | 351.72 | 63.16 | 148.93 | 280.09 | 0.16 | 8710.21 |
| SmallAreaLowGrayLevelEmphasis | 0.08 | 0.09 | 0.02 | 0.04 | 0.10 | 0.00 | 0.68 |
| ZoneEntropy | 4.52 | 1.36 | 3.55 | 4.76 | 5.57 | 0.92 | 7.90 |
| ZonePercentage | 0.67 | 0.19 | 0.57 | 0.68 | 0.80 | 0.01 | 1.00 |
| ZoneVariance | 236.47 | 10641.88 | 0.24 | 0.78 | 2.24 | 0.00 | 1226340.91 |
| **Neighbouring Gray Tone Difference Matrix (NGTDM)** | | | | | | | |
| Busyness | 0.18 | 0.34 | 0.06 | 0.10 | 0.19 | 0.00 | 14.98 |
| Coarseness | 0.09 | 0.11 | 0.03 | 0.06 | 0.12 | 0.00 | 1.50 |
| Complexity | 1859.31 | 5011.82 | 243.90 | 850.51 | 1871.17 | 0.22 | 173314.58 |
| Contrast | 11.71 | 132.30 | 0.37 | 0.64 | 1.59 | 0.01 | 8884.17 |
| Strength | 16.05 | 16.10 | 7.00 | 12.41 | 19.69 | 0.03 | 187.81 |
| **Shape (3D)** | | | | | | | |
| Elongation | 0.73 | 0.15 | 0.63 | 0.74 | 0.84 | 0.02 | 1.00 |
| Flatness | 0.52 | 0.18 | 0.43 | 0.54 | 0.64 | 0.00 | 1.00 |
| LeastAxisLength | 4.23 | 3.50 | 2.06 | 3.61 | 5.24 | 0.00 | 51.43 |
| MajorAxisLength | 8.22 | 7.20 | 4.18 | 6.55 | 9.40 | 2.00 | 250.32 |
| Maximum2DDiameterColumn | 8.45 | 7.62 | 4.12 | 6.71 | 10.00 | 1.00 | 259.01 |
| Maximum2DDiameterRow | 8.40 | 7.45 | 4.12 | 6.71 | 10.00 | 1.00 | 259.02 |
| Maximum2DDiameterSlice | 8.05 | 6.96 | 4.12 | 6.32 | 9.22 | 1.00 | 110.55 |
| Maximum3DDiameter | 9.69 | 8.74 | 4.58 | 7.81 | 11.18 | 2.24 | 259.03 |
| MeshVolume | 513.89 | 3889.98 | 14.79 | 66.29 | 191.17 | 0.83 | 237030.42 |
| MinorAxisLength | 5.82 | 4.66 | 2.97 | 4.84 | 6.82 | 0.83 | 77.13 |
| Sphericity | 0.77 | 0.09 | 0.72 | 0.78 | 0.83 | 0.13 | 0.94 |
| SurfaceArea | 288.16 | 918.09 | 35.68 | 102.58 | 215.23 | 6.22 | 31775.90 |
| SurfaceVolumeRatio | 1.91 | 1.11 | 1.13 | 1.55 | 2.45 | 0.13 | 8.80 |
| VoxelVolume | 522.67 | 3895.85 | 18.00 | 73.00 | 201.00 | 2.06 | 237180.00 |

Abbreviations: P25, $25^{th}$ percentile; P75, $75^{th}$ percentile; SD, standard deviation.

**Supplemental Table 4.** Set of 642 radiomics features retained after the filtering steps described in the Supplementary Methods and in Supplemental Figure 2.

| Class | Features |
|---|---|
| First Order | original_firstorder_10Percentile, original_firstorder_Kurtosis, original_firstorder_Maximum, original_firstorder_MeanAbsoluteDeviation, original_firstorder_Mean, original_firstorder_Minimum, original_firstorder_Range, original_firstorder_Skewness, wavelet-LLH_firstorder_10Percentile, wavelet-LLH_firstorder_Maximum, wavelet-LLH_firstorder_MeanAbsoluteDeviation, wavelet-LLH_firstorder_Mean, wavelet-LHL_firstorder_10Percentile, wavelet-LHL_firstorder_Kurtosis, wavelet-LHL_firstorder_Maximum, wavelet-LHL_firstorder_MeanAbsoluteDeviation, wavelet-LHL_firstorder_Mean, wavelet-LHL_firstorder_Minimum, wavelet-LHL_firstorder_Range, wavelet-LHL_firstorder_Skewness, wavelet-LHH_firstorder_10Percentile, wavelet-LHH_firstorder_Maximum, wavelet-LHH_firstorder_MeanAbsoluteDeviation, wavelet-LHH_firstorder_Mean, wavelet-LHH_firstorder_Minimum, wavelet-LHH_firstorder_Range, wavelet-HLL_firstorder_10Percentile, wavelet-HLL_firstorder_Kurtosis, wavelet-HLL_firstorder_Maximum, wavelet-HLL_firstorder_MeanAbsoluteDeviation, wavelet-HLL_firstorder_Mean, wavelet-HLL_firstorder_Minimum, wavelet-HLL_firstorder_Range, wavelet-HLL_firstorder_Skewness, wavelet-HLH_firstorder_10Percentile, wavelet-HLH_firstorder_Kurtosis, wavelet-HLH_firstorder_Maximum, wavelet-HLH_firstorder_MeanAbsoluteDeviation, wavelet-HLH_firstorder_Minimum, wavelet-HLH_firstorder_Range, wavelet-HHL_firstorder_Kurtosis, wavelet-HHL_firstorder_Maximum, wavelet-HHL_firstorder_MeanAbsoluteDeviation, wavelet-HHL_firstorder_Mean, wavelet-HHL_firstorder_Minimum, wavelet-HHL_firstorder_Range, wavelet-HHH_firstorder_10Percentile, wavelet-HHH_firstorder_Maximum, wavelet-HHH_firstorder_MeanAbsoluteDeviation, wavelet-HHH_firstorder_Minimum, wavelet-HHH_firstorder_Range, wavelet-LLL_firstorder_10Percentile, wavelet-LLL_firstorder_Kurtosis, wavelet-LLL_firstorder_Maximum, wavelet-LLL_firstorder_MeanAbsoluteDeviation, wavelet-LLL_firstorder_Mean, wavelet-LLL_firstorder_Minimum, wavelet-LLL_firstorder_Range, wavelet-LLL_firstorder_Skewness, log-sigma-1-0-mm-3D_firstorder_10Percentile, log-sigma-1-0-mm-3D_firstorder_Kurtosis, log-sigma-1-0-mm-3D_firstorder_Maximum, log-sigma-1-0-mm-3D_firstorder_MeanAbsoluteDeviation, log-sigma-1-0-mm-3D_firstorder_Mean, log-sigma-1-0-mm-3D_firstorder_Minimum, log-sigma-1-0-mm-3D_firstorder_Range, log-sigma-2-0-mm-3D_firstorder_10Percentile, log-sigma-2-0-mm-3D_firstorder_Kurtosis, log-sigma-2-0-mm-3D_firstorder_Maximum, log-sigma-2-0-mm-3D_firstorder_MeanAbsoluteDeviation, log-sigma-2-0-mm-3D_firstorder_Minimum, log-sigma-2-0-mm-3D_firstorder_Range, log-sigma-2-0-mm-3D_firstorder_Skewness, log-sigma-3-0-mm-3D_firstorder_10Percentile, log-sigma-3-0-mm-3D_firstorder_Kurtosis, log-sigma-3-0-mm-3D_firstorder_Maximum, log-sigma-3-0-mm-3D_firstorder_MeanAbsoluteDeviation, log-sigma-3-0-mm-3D_firstorder_Mean, log-sigma-3-0-mm-3D_firstorder_Minimum, log-sigma-3-0-mm-3D_firstorder_Range, log-sigma-3-0-mm-3D_firstorder_Skewness, log-sigma-4-0-mm-3D_firstorder_10Percentile, log-sigma-4-0-mm-3D_firstorder_Kurtosis, log-sigma-4-0-mm-3D_firstorder_Maximum, log-sigma-4-0-mm-3D_firstorder_MeanAbsoluteDeviation, log-sigma-4-0-mm-3D_firstorder_Mean, log-sigma-4-0-mm-3D_firstorder_Minimum, log-sigma-4-0-mm-3D_firstorder_Range, log-sigma-5-0-mm-3D_firstorder_10Percentile, log-sigma-5-0-mm-3D_firstorder_MeanAbsoluteDeviation, log-sigma-5-0-mm-3D_firstorder_Mean, log-sigma-5-0-mm-3D_firstorder_Minimum, log-sigma-5-0-mm-3D_firstorder_Range, log-sigma-5-0-mm-3D_firstorder_Skewness, square_firstorder_10Percentile, square_firstorder_Kurtosis, square_firstorder_Maximum, square_firstorder_Mean, square_firstorder_Minimum, square_firstorder_Range, square_firstorder_Skewness, squareroot_firstorder_Kurtosis, squareroot_firstorder_Maximum, squareroot_firstorder_MeanAbsoluteDeviation, squareroot_firstorder_Mean, squareroot_firstorder_Minimum, squareroot_firstorder_Range, logarithm_firstorder_Kurtosis, logarithm_firstorder_Maximum, logarithm_firstorder_MeanAbsoluteDeviation, logarithm_firstorder_Mean, logarithm_firstorder_Minimum, logarithm_firstorder_Range, logarithm_firstorder_Skewness, exponential_firstorder_Kurtosis, exponential_firstorder_Maximum, exponential_firstorder_Mean, exponential_firstorder_Minimum, exponential_firstorder_Range, exponential_firstorder_Skewness, gradient_firstorder_10Percentile, gradient_firstorder_Kurtosis, gradient_firstorder_Maximum, gradient_firstorder_MeanAbsoluteDeviation, gradient_firstorder_Mean, gradient_firstorder_Minimum, gradient_firstorder_Range, gradient_firstorder_Skewness, lbp-3D-m1_firstorder_10Percentile, lbp-3D-m1_firstorder_Maximum, lbp-3D-m1_firstorder_MeanAbsoluteDeviation, lbp-3D-m1_firstorder_Mean, lbp-3D-m1_firstorder_Minimum, lbp-3D-m1_firstorder_Range, lbp-3D-m1_firstorder_Skewness, lbp-3D-m2_firstorder_10Percentile, lbp-3D-m2_firstorder_Maximum, lbp-3D-m2_firstorder_MeanAbsoluteDeviation, lbp-3D-m2_firstorder_Mean, lbp-3D-m2_firstorder_Minimum, lbp-3D-m2_firstorder_Range, lbp-3D-k_firstorder_10Percentile, lbp-3D-k_firstorder_Kurtosis, lbp-3D-k_firstorder_Maximum, lbp-3D-k_firstorder_MeanAbsoluteDeviation, lbp-3D-k_firstorder_Mean, lbp-3D-k_firstorder_Minimum, lbp-3D-k_firstorder_Range, lbp-3D-k_firstorder_Skewness |

| | |
|---|---|
| GLCM | original_glcm_ClusterShade, original_glcm_ClusterTendency, original_glcm_DifferenceAverage, original_glcm_Idm, original_glcm_Idn, original_glcm_Imc1, original_glcm_Imc2, original_glcm_InverseVariance, original_glcm_MCC, wavelet-LLH_glcm_ClusterProminence, wavelet-LLH_glcm_ClusterTendency, wavelet-LLH_glcm_DifferenceAverage, wavelet-LLH_glcm_Idm, wavelet-LLH_glcm_Idn, wavelet-LLH_glcm_Imc1, wavelet-LLH_glcm_Imc2, wavelet-LLH_glcm_InverseVariance, wavelet-LLH_glcm_MCC, wavelet-LHL_glcm_ClusterProminence, wavelet-LHL_glcm_ClusterTendency, wavelet-LHL_glcm_DifferenceAverage, wavelet-LHL_glcm_Idm, wavelet-LHL_glcm_Idn, wavelet-LHL_glcm_Imc1, wavelet-LHL_glcm_Imc2, wavelet-LHL_glcm_InverseVariance, wavelet-LHL_glcm_MCC, wavelet-LHH_glcm_ClusterTendency, wavelet-LHH_glcm_DifferenceAverage, wavelet-LHH_glcm_Idm, wavelet-LHH_glcm_Idn, wavelet-LHH_glcm_Imc1, wavelet-LHH_glcm_Imc2, wavelet-LHH_glcm_InverseVariance, wavelet-LHH_glcm_MCC, wavelet-HLL_glcm_ClusterProminence, wavelet-HLL_glcm_ClusterTendency, wavelet-HLL_glcm_DifferenceAverage, wavelet-HLL_glcm_Idm, wavelet-HLL_glcm_Idn, wavelet-HLL_glcm_Imc1, wavelet-HLL_glcm_Imc2, wavelet-HLL_glcm_InverseVariance, wavelet-HLL_glcm_MCC, wavelet-HLH_glcm_ClusterProminence, wavelet-HLH_glcm_ClusterTendency, wavelet-HLH_glcm_DifferenceAverage, wavelet-HLH_glcm_Idm, wavelet-HLH_glcm_Idn, wavelet-HLH_glcm_Imc1, wavelet-HLH_glcm_Imc2, wavelet-HLH_glcm_InverseVariance, wavelet-HLH_glcm_MCC, wavelet-HHL_glcm_DifferenceAverage, wavelet-HHL_glcm_Idm, wavelet-HHL_glcm_Idn, wavelet-HHL_glcm_Imc1, wavelet-HHL_glcm_Imc2, wavelet-HHL_glcm_InverseVariance, wavelet-HHL_glcm_MCC, wavelet-HHH_glcm_DifferenceAverage, wavelet-HHH_glcm_Idm, wavelet-HHH_glcm_Idn, wavelet-HHH_glcm_Imc1, wavelet-HHH_glcm_Imc2, wavelet-HHH_glcm_InverseVariance, wavelet-HHH_glcm_MCC, wavelet-LLL_glcm_ClusterShade, wavelet-LLL_glcm_ClusterTendency, wavelet-LLL_glcm_DifferenceAverage, wavelet-LLL_glcm_Idm, wavelet-LLL_glcm_Idn, wavelet-LLL_glcm_Imc1, wavelet-LLL_glcm_Imc2, wavelet-LLL_glcm_InverseVariance, wavelet-LLL_glcm_MCC, log-sigma-1-0-mm-3D_glcm_DifferenceAverage, log-sigma-1-0-mm-3D_glcm_Idm, log-sigma-1-0-mm-3D_glcm_Idn, log-sigma-1-0-mm-3D_glcm_Imc1, log-sigma-1-0-mm-3D_glcm_Imc2, log-sigma-1-0-mm-3D_glcm_InverseVariance, log-sigma-1-0-mm-3D_glcm_MCC, log-sigma-2-0-mm-3D_glcm_ClusterTendency, log-sigma-2-0-mm-3D_glcm_Idn, log-sigma-2-0-mm-3D_glcm_Imc1, log-sigma-2-0-mm-3D_glcm_Imc2, log-sigma-3-0-mm-3D_glcm_ClusterProminence, log-sigma-3-0-mm-3D_glcm_ClusterTendency, log-sigma-3-0-mm-3D_glcm_DifferenceAverage, log-sigma-3-0-mm-3D_glcm_Idm, log-sigma-3-0-mm-3D_glcm_Idn, log-sigma-3-0-mm-3D_glcm_Imc1, log-sigma-3-0-mm-3D_glcm_Imc2, log-sigma-3-0-mm-3D_glcm_InverseVariance, log-sigma-3-0-mm-3D_glcm_MCC, log-sigma-4-0-mm-3D_glcm_ClusterProminence, log-sigma-4-0-mm-3D_glcm_ClusterShade, log-sigma-4-0-mm-3D_glcm_ClusterTendency, log-sigma-4-0-mm-3D_glcm_DifferenceAverage, log-sigma-4-0-mm-3D_glcm_Idm, log-sigma-4-0-mm-3D_glcm_Idn, log-sigma-4-0-mm-3D_glcm_Imc1, log-sigma-4-0-mm-3D_glcm_Imc2, log-sigma-4-0-mm-3D_glcm_InverseVariance, log-sigma-4-0-mm-3D_glcm_MCC, log-sigma-5-0-mm-3D_glcm_ClusterProminence, log-sigma-5-0-mm-3D_glcm_ClusterShade, log-sigma-5-0-mm-3D_glcm_ClusterTendency, log-sigma-5-0-mm-3D_glcm_DifferenceAverage, log-sigma-5-0-mm-3D_glcm_Idm, log-sigma-5-0-mm-3D_glcm_Idn, log-sigma-5-0-mm-3D_glcm_Imc1, log-sigma-5-0-mm-3D_glcm_Imc2, log-sigma-5-0-mm-3D_glcm_InverseVariance, log-sigma-5-0-mm-3D_glcm_MCC, square_glcm_DifferenceAverage, square_glcm_Idm, square_glcm_Idn, square_glcm_Imc1, square_glcm_Imc2, square_glcm_InverseVariance, square_glcm_MCC, squareroot_glcm_ClusterTendency, squareroot_glcm_DifferenceAverage, squareroot_glcm_Idn, squareroot_glcm_Imc1, squareroot_glcm_Imc2, squareroot_glcm_MCC, logarithm_glcm_ClusterProminence, logarithm_glcm_ClusterTendency, logarithm_glcm_Idm, logarithm_glcm_Idn, logarithm_glcm_Imc1, logarithm_glcm_InverseVariance, logarithm_glcm_MCC, exponential_glcm_Imc1, exponential_glcm_Imc2, exponential_glcm_InverseVariance, gradient_glcm_ClusterShade, gradient_glcm_ClusterTendency, gradient_glcm_DifferenceAverage, gradient_glcm_Idm, gradient_glcm_Idn, gradient_glcm_Imc1, gradient_glcm_Imc2, gradient_glcm_InverseVariance, gradient_glcm_MCC, lbp-3D-k_glcm_ClusterProminence, lbp-3D-k_glcm_ClusterShade, lbp-3D-k_glcm_ClusterTendency, lbp-3D-k_glcm_DifferenceAverage, lbp-3D-k_glcm_Idm, lbp-3D-k_glcm_Idn, lbp-3D-k_glcm_Imc1, lbp-3D-k_glcm_InverseVariance, lbp-3D-k_glcm_MCC |
| GLDM | original_gldm_DependenceNonUniformityNormalized, original_gldm_LargeDependenceEmphasis, original_gldm_LargeDependenceHighGrayLevelEmphasis, original_gldm_LargeDependenceLowGrayLevelEmphasis, wavelet-LLH_gldm_DependenceNonUniformityNormalized, wavelet-LLH_gldm_LargeDependenceEmphasis, wavelet-LLH_gldm_LargeDependenceHighGrayLevelEmphasis, wavelet-LHL_gldm_DependenceNonUniformityNormalized, wavelet-LHL_gldm_LargeDependenceEmphasis, wavelet-LHL_gldm_LargeDependenceHighGrayLevelEmphasis, wavelet-LHL_gldm_LargeDependenceLowGrayLevelEmphasis, wavelet-LHH_gldm_DependenceNonUniformityNormalized, wavelet-LHH_gldm_LargeDependenceEmphasis, wavelet-LHH_gldm_LargeDependenceHighGrayLevelEmphasis, wavelet- |

| | |
|---|---|
| | LHH_gldm_LargeDependenceLowGrayLevelEmphasis, wavelet-HLL_gldm_DependenceNonUniformityNormalized, wavelet-HLL_gldm_LargeDependenceEmphasis, wavelet-HLL_gldm_LargeDependenceHighGrayLevelEmphasis, wavelet-HLL_gldm_LargeDependenceLowGrayLevelEmphasis, wavelet-HLH_gldm_DependenceNonUniformityNormalized, wavelet-HLH_gldm_LargeDependenceEmphasis, wavelet-HLH_gldm_LargeDependenceHighGrayLevelEmphasis, wavelet-HHL_gldm_DependenceNonUniformityNormalized, wavelet-HHL_gldm_LargeDependenceEmphasis, wavelet-HHL_gldm_LargeDependenceHighGrayLevelEmphasis, wavelet-HHL_gldm_LargeDependenceLowGrayLevelEmphasis, wavelet-HHH_gldm_DependenceNonUniformityNormalized, wavelet-HHH_gldm_LargeDependenceEmphasis, wavelet-HHH_gldm_LargeDependenceHighGrayLevelEmphasis, wavelet-HHH_gldm_LargeDependenceLowGrayLevelEmphasis, wavelet-LLL_gldm_DependenceNonUniformityNormalized, wavelet-LLL_gldm_LargeDependenceEmphasis, wavelet-LLL_gldm_LargeDependenceHighGrayLevelEmphasis, wavelet-LLL_gldm_LargeDependenceLowGrayLevelEmphasis, log-sigma-1-0-mm-3D_gldm_DependenceNonUniformityNormalized, log-sigma-1-0-mm-3D_gldm_LargeDependenceEmphasis, log-sigma-1-0-mm-3D_gldm_LargeDependenceHighGrayLevelEmphasis, log-sigma-2-0-mm-3D_gldm_DependenceNonUniformityNormalized, log-sigma-2-0-mm-3D_gldm_LargeDependenceEmphasis, log-sigma-2-0-mm-3D_gldm_LargeDependenceHighGrayLevelEmphasis, log-sigma-2-0-mm-3D_gldm_LargeDependenceLowGrayLevelEmphasis, log-sigma-3-0-mm-3D_gldm_DependenceNonUniformityNormalized, log-sigma-3-0-mm-3D_gldm_LargeDependenceEmphasis, log-sigma-3-0-mm-3D_gldm_LargeDependenceHighGrayLevelEmphasis, log-sigma-3-0-mm-3D_gldm_LargeDependenceLowGrayLevelEmphasis, log-sigma-4-0-mm-3D_gldm_DependenceNonUniformityNormalized, log-sigma-4-0-mm-3D_gldm_LargeDependenceHighGrayLevelEmphasis, log-sigma-4-0-mm-3D_gldm_LargeDependenceLowGrayLevelEmphasis, log-sigma-5-0-mm-3D_gldm_DependenceNonUniformityNormalized, log-sigma-5-0-mm-3D_gldm_LargeDependenceHighGrayLevelEmphasis, log-sigma-5-0-mm-3D_gldm_LargeDependenceLowGrayLevelEmphasis, square_gldm_DependenceNonUniformityNormalized, square_gldm_LargeDependenceEmphasis, square_gldm_LargeDependenceHighGrayLevelEmphasis, square_gldm_LargeDependenceLowGrayLevelEmphasis, squareroot_gldm_DependenceNonUniformityNormalized, squareroot_gldm_LargeDependenceEmphasis, squareroot_gldm_LargeDependenceHighGrayLevelEmphasis, logarithm_gldm_DependenceNonUniformityNormalized, logarithm_gldm_LargeDependenceEmphasis, logarithm_gldm_LargeDependenceHighGrayLevelEmphasis, logarithm_gldm_LargeDependenceLowGrayLevelEmphasis, exponential_gldm_LargeDependenceEmphasis, exponential_gldm_LargeDependenceHighGrayLevelEmphasis, exponential_gldm_LargeDependenceLowGrayLevelEmphasis, gradient_gldm_DependenceNonUniformityNormalized, gradient_gldm_LargeDependenceEmphasis, gradient_gldm_LargeDependenceHighGrayLevelEmphasis, gradient_gldm_LargeDependenceLowGrayLevelEmphasis, lbp-3D-m1_gldm_LargeDependenceEmphasis, lbp-3D-m1_gldm_LargeDependenceHighGrayLevelEmphasis, lbp-3D-m1_gldm_LargeDependenceLowGrayLevelEmphasis, lbp-3D-m2_gldm_LargeDependenceEmphasis, lbp-3D-m2_gldm_LargeDependenceHighGrayLevelEmphasis, lbp-3D-m2_gldm_LargeDependenceLowGrayLevelEmphasis, lbp-3D-k_gldm_DependenceNonUniformityNormalized, lbp-3D-k_gldm_LargeDependenceEmphasis, lbp-3D-k_gldm_LargeDependenceHighGrayLevelEmphasis, lbp-3D-k_gldm_LargeDependenceLowGrayLevelEmphasis |
| GLRLM | original_glrlm_GrayLevelNonUniformity, original_glrlm_LongRunLowGrayLevelEmphasis, original_glrlm_RunEntropy, original_glrlm_RunPercentage, original_glrlm_ShortRunLowGrayLevelEmphasis, wavelet-LLH_glrlm_GrayLevelNonUniformity, wavelet-LLH_glrlm_LongRunLowGrayLevelEmphasis, wavelet-LLH_glrlm_RunEntropy, wavelet-LLH_glrlm_RunPercentage, wavelet-LLH_glrlm_RunVariance, wavelet-LLH_glrlm_ShortRunLowGrayLevelEmphasis, wavelet-LHL_glrlm_GrayLevelNonUniformity, wavelet-LHL_glrlm_LongRunLowGrayLevelEmphasis, wavelet-LHL_glrlm_RunEntropy, wavelet-LHL_glrlm_RunPercentage, wavelet-LHL_glrlm_ShortRunLowGrayLevelEmphasis, wavelet-LHH_glrlm_GrayLevelNonUniformity, wavelet-LHH_glrlm_LongRunLowGrayLevelEmphasis, wavelet-LHH_glrlm_RunEntropy, wavelet-LHH_glrlm_RunPercentage, wavelet-LHH_glrlm_RunVariance, wavelet-LHH_glrlm_ShortRunLowGrayLevelEmphasis, wavelet-HLL_glrlm_GrayLevelNonUniformity, wavelet-HLL_glrlm_LongRunLowGrayLevelEmphasis, wavelet-HLL_glrlm_RunEntropy, wavelet-HLL_glrlm_RunPercentage, wavelet-HLL_glrlm_ShortRunLowGrayLevelEmphasis, wavelet-HLH_glrlm_GrayLevelNonUniformity, wavelet-HLH_glrlm_LongRunLowGrayLevelEmphasis, wavelet-HLH_glrlm_RunEntropy, wavelet-HLH_glrlm_RunPercentage, wavelet-HLH_glrlm_RunVariance, wavelet-HLH_glrlm_ShortRunLowGrayLevelEmphasis, wavelet-HHL_glrlm_GrayLevelNonUniformity, wavelet-HHL_glrlm_LongRunLowGrayLevelEmphasis, wavelet-HHL_glrlm_RunEntropy, wavelet-HHL_glrlm_RunPercentage, wavelet- |

| | |
|---|---|
| | HHL_glrlm_ShortRunLowGrayLevelEmphasis, wavelet-HHH_glrlm_GrayLevelNonUniformity, wavelet-HHH_glrlm_LongRunLowGrayLevelEmphasis, wavelet-HHH_glrlm_RunEntropy, wavelet-HHH_glrlm_RunPercentage, wavelet-HHH_glrlm_RunVariance, wavelet-HHH_glrlm_ShortRunLowGrayLevelEmphasis, wavelet-LLL_glrlm_GrayLevelNonUniformity, wavelet-LLL_glrlm_LongRunLowGrayLevelEmphasis, wavelet-LLL_glrlm_RunEntropy, wavelet-LLL_glrlm_RunPercentage, wavelet-LLL_glrlm_RunVariance, wavelet-LLL_glrlm_ShortRunLowGrayLevelEmphasis, log-sigma-1-0-mm-3D_glrlm_GrayLevelNonUniformity, log-sigma-1-0-mm-3D_glrlm_LongRunLowGrayLevelEmphasis, log-sigma-1-0-mm-3D_glrlm_RunEntropy, log-sigma-1-0-mm-3D_glrlm_RunPercentage, log-sigma-1-0-mm-3D_glrlm_ShortRunLowGrayLevelEmphasis, log-sigma-2-0-mm-3D_glrlm_GrayLevelNonUniformity, log-sigma-2-0-mm-3D_glrlm_LongRunLowGrayLevelEmphasis, log-sigma-2-0-mm-3D_glrlm_RunEntropy, log-sigma-2-0-mm-3D_glrlm_RunPercentage, log-sigma-2-0-mm-3D_glrlm_ShortRunLowGrayLevelEmphasis, log-sigma-3-0-mm-3D_glrlm_GrayLevelNonUniformity, log-sigma-3-0-mm-3D_glrlm_LongRunLowGrayLevelEmphasis, log-sigma-3-0-mm-3D_glrlm_RunEntropy, log-sigma-3-0-mm-3D_glrlm_RunPercentage, log-sigma-3-0-mm-3D_glrlm_ShortRunLowGrayLevelEmphasis, log-sigma-4-0-mm-3D_glrlm_GrayLevelNonUniformity, log-sigma-4-0-mm-3D_glrlm_LongRunLowGrayLevelEmphasis, log-sigma-4-0-mm-3D_glrlm_RunEntropy, log-sigma-4-0-mm-3D_glrlm_ShortRunLowGrayLevelEmphasis, log-sigma-5-0-mm-3D_glrlm_GrayLevelNonUniformity, log-sigma-5-0-mm-3D_glrlm_LongRunLowGrayLevelEmphasis, log-sigma-5-0-mm-3D_glrlm_RunEntropy, log-sigma-5-0-mm-3D_glrlm_ShortRunLowGrayLevelEmphasis, square_glrlm_GrayLevelNonUniformity, square_glrlm_LongRunLowGrayLevelEmphasis, square_glrlm_RunEntropy, square_glrlm_RunPercentage, square_glrlm_RunVariance, square_glrlm_ShortRunLowGrayLevelEmphasis, squareroot_glrlm_GrayLevelNonUniformity, squareroot_glrlm_LongRunLowGrayLevelEmphasis, squareroot_glrlm_RunEntropy, squareroot_glrlm_RunPercentage, squareroot_glrlm_ShortRunLowGrayLevelEmphasis, logarithm_glrlm_GrayLevelNonUniformity, logarithm_glrlm_LongRunLowGrayLevelEmphasis, logarithm_glrlm_RunEntropy, logarithm_glrlm_RunPercentage, logarithm_glrlm_ShortRunLowGrayLevelEmphasis, exponential_glrlm_GrayLevelNonUniformity, exponential_glrlm_LongRunLowGrayLevelEmphasis, exponential_glrlm_RunEntropy, exponential_glrlm_RunPercentage, exponential_glrlm_RunVariance, exponential_glrlm_ShortRunLowGrayLevelEmphasis, gradient_glrlm_GrayLevelNonUniformity, gradient_glrlm_LongRunLowGrayLevelEmphasis, gradient_glrlm_RunEntropy, gradient_glrlm_RunPercentage, gradient_glrlm_ShortRunLowGrayLevelEmphasis, lbp-3D-m1_glrlm_GrayLevelNonUniformity, lbp-3D-m1_glrlm_LongRunLowGrayLevelEmphasis, lbp-3D-m1_glrlm_RunEntropy, lbp-3D-m1_glrlm_RunPercentage, lbp-3D-m1_glrlm_RunVariance, lbp-3D-m1_glrlm_ShortRunLowGrayLevelEmphasis, lbp-3D-m2_glrlm_GrayLevelNonUniformity, lbp-3D-m2_glrlm_LongRunLowGrayLevelEmphasis, lbp-3D-m2_glrlm_RunEntropy, lbp-3D-m2_glrlm_RunPercentage, lbp-3D-m2_glrlm_RunVariance, lbp-3D-m2_glrlm_ShortRunLowGrayLevelEmphasis, lbp-3D-k_glrlm_GrayLevelNonUniformity, lbp-3D-k_glrlm_LongRunLowGrayLevelEmphasis, lbp-3D-k_glrlm_RunEntropy, lbp-3D-k_glrlm_RunPercentage, lbp-3D-k_glrlm_RunVariance, lbp-3D-k_glrlm_ShortRunLowGrayLevelEmphasis |
| GLSZM | original_glszm_GrayLevelNonUniformity, original_glszm_HighGrayLevelZoneEmphasis, original_glszm_ZonePercentage, wavelet-LLH_glszm_GrayLevelNonUniformity, wavelet-LLH_glszm_ZonePercentage, wavelet-LHL_glszm_GrayLevelNonUniformity, wavelet-LHL_glszm_HighGrayLevelZoneEmphasis, wavelet-LHL_glszm_ZonePercentage, wavelet-LHH_glszm_GrayLevelNonUniformity, wavelet-LHH_glszm_HighGrayLevelZoneEmphasis, wavelet-LHH_glszm_ZonePercentage, wavelet-HLL_glszm_GrayLevelNonUniformity, wavelet-HLL_glszm_HighGrayLevelZoneEmphasis, wavelet-HLL_glszm_ZonePercentage, wavelet-HLH_glszm_GrayLevelNonUniformity, wavelet-HLH_glszm_HighGrayLevelZoneEmphasis, wavelet-HLH_glszm_ZonePercentage, wavelet-HHL_glszm_GrayLevelNonUniformity, wavelet-HHL_glszm_HighGrayLevelZoneEmphasis, wavelet-HHL_glszm_ZonePercentage, wavelet-HHH_glszm_GrayLevelNonUniformity, wavelet-HHH_glszm_HighGrayLevelZoneEmphasis, wavelet-HHH_glszm_ZonePercentage, wavelet-LLL_glszm_GrayLevelNonUniformity, wavelet-LLL_glszm_HighGrayLevelZoneEmphasis, wavelet-LLL_glszm_ZonePercentage, log-sigma-1-0-mm-3D_glszm_GrayLevelNonUniformity, log-sigma-1-0-mm-3D_glszm_HighGrayLevelZoneEmphasis, log-sigma-1-0-mm-3D_glszm_ZonePercentage, log-sigma-2-0-mm-3D_glszm_GrayLevelNonUniformity, log-sigma-2-0-mm-3D_glszm_HighGrayLevelZoneEmphasis, log-sigma-2-0-mm-3D_glszm_ZonePercentage, log-sigma-3-0-mm-3D_glszm_GrayLevelNonUniformity, log-sigma-3-0-mm-3D_glszm_HighGrayLevelZoneEmphasis, log-sigma-3-0-mm-3D_glszm_ZonePercentage, log-sigma-4-0-mm-3D_glszm_GrayLevelNonUniformity, log-sigma-4-0-mm-3D_glszm_HighGrayLevelZoneEmphasis, log-sigma-4-0-mm-3D_glszm_ZonePercentage, log-sigma-5-0-mm- |

| | |
|---|---|
| | 3D_glszm_GrayLevelNonUniformity, log-sigma-5-0-mm-3D_glszm_HighGrayLevelZoneEmphasis, log-sigma-5-0-mm-3D_glszm_ZonePercentage, square_glszm_GrayLevelNonUniformity, square_glszm_ZonePercentage, squareroot_glszm_GrayLevelNonUniformity, squareroot_glszm_HighGrayLevelZoneEmphasis, squareroot_glszm_ZonePercentage, logarithm_glszm_GrayLevelNonUniformity, logarithm_glszm_HighGrayLevelZoneEmphasis, logarithm_glszm_ZonePercentage, exponential_glszm_GrayLevelNonUniformity, exponential_glszm_ZonePercentage, gradient_glszm_GrayLevelNonUniformity, gradient_glszm_ZonePercentage, lbp-3D-m1_glszm_ZonePercentage, lbp-3D-m2_glszm_ZonePercentage, lbp-3D-k_glszm_GrayLevelNonUniformity, lbp-3D-k_glszm_HighGrayLevelZoneEmphasis, lbp-3D-k_glszm_ZonePercentage |
| NGTDM | original_ngtdm_Busyness, original_ngtdm_Complexity, original_ngtdm_Contrast, original_ngtdm_Strength, wavelet-LLH_ngtdm_Busyness, wavelet-LLH_ngtdm_Contrast, wavelet-LLH_ngtdm_Strength, wavelet-LHL_ngtdm_Busyness, wavelet-LHL_ngtdm_Complexity, wavelet-LHL_ngtdm_Contrast, wavelet-LHL_ngtdm_Strength, wavelet-LHH_ngtdm_Busyness, wavelet-LHH_ngtdm_Complexity, wavelet-LHH_ngtdm_Contrast, wavelet-LHH_ngtdm_Strength, wavelet-HLL_ngtdm_Busyness, wavelet-HLL_ngtdm_Complexity, wavelet-HLL_ngtdm_Contrast, wavelet-HLL_ngtdm_Strength, wavelet-HLH_ngtdm_Busyness, wavelet-HLH_ngtdm_Complexity, wavelet-HLH_ngtdm_Contrast, wavelet-HLH_ngtdm_Strength, wavelet-HHL_ngtdm_Busyness, wavelet-HHL_ngtdm_Complexity, wavelet-HHL_ngtdm_Contrast, wavelet-HHL_ngtdm_Strength, wavelet-HHH_ngtdm_Busyness, wavelet-HHH_ngtdm_Complexity, wavelet-HHH_ngtdm_Contrast, wavelet-HHH_ngtdm_Strength, wavelet-LLL_ngtdm_Complexity, wavelet-LLL_ngtdm_Contrast, wavelet-LLL_ngtdm_Strength, log-sigma-1-0-mm-3D_ngtdm_Busyness, log-sigma-1-0-mm-3D_ngtdm_Contrast, log-sigma-1-0-mm-3D_ngtdm_Strength, log-sigma-2-0-mm-3D_ngtdm_Busyness, log-sigma-2-0-mm-3D_ngtdm_Complexity, log-sigma-2-0-mm-3D_ngtdm_Contrast, log-sigma-2-0-mm-3D_ngtdm_Strength, log-sigma-3-0-mm-3D_ngtdm_Complexity, log-sigma-3-0-mm-3D_ngtdm_Contrast, log-sigma-3-0-mm-3D_ngtdm_Strength, log-sigma-4-0-mm-3D_ngtdm_Complexity, log-sigma-4-0-mm-3D_ngtdm_Contrast, log-sigma-4-0-mm-3D_ngtdm_Strength, log-sigma-5-0-mm-3D_ngtdm_Complexity, log-sigma-5-0-mm-3D_ngtdm_Contrast, log-sigma-5-0-mm-3D_ngtdm_Strength, square_ngtdm_Busyness, square_ngtdm_Contrast, square_ngtdm_Strength, squareroot_ngtdm_Busyness, squareroot_ngtdm_Complexity, squareroot_ngtdm_Contrast, squareroot_ngtdm_Strength, logarithm_ngtdm_Complexity, logarithm_ngtdm_Contrast, logarithm_ngtdm_Strength, exponential_ngtdm_Strength, gradient_ngtdm_Busyness, gradient_ngtdm_Complexity, gradient_ngtdm_Contrast, gradient_ngtdm_Strength, lbp-3D-k_ngtdm_Busyness, lbp-3D-k_ngtdm_Complexity, lbp-3D-k_ngtdm_Strength, wavelet-LLH_ngtdm_Coarseness, wavelet-LHL_ngtdm_Coarseness, wavelet-LHH_ngtdm_Coarseness, wavelet-HLL_ngtdm_Coarseness, wavelet-HLH_ngtdm_Coarseness, wavelet-HHL_ngtdm_Coarseness, wavelet-HHH_ngtdm_Coarseness, wavelet-LLL_ngtdm_Coarseness, original_ngtdm_Coarseness |
| Shape (3D) | original_shape_Elongation, original_shape_Flatness, original_shape_LeastAxisLength, original_shape_Sphericity |

**Supplemental Table 5.** Comparison of model performance between our radiomics model and the established Brock Model. We provide point estimates and percentile-based bootstrap confidence intervals for the discrimination and calibration metrics.

| | Observed number of cancers (95% CI) | Expected number of cancers (95% CI) | Expected / Observed (95% CI) | Expected – Observed (95% CI) | Area under the curve (95% CI) |
|---|---|---|---|---|---|
| Radiomics [1] | 3181.68 (2616.71, 3806.13) | 3251.08 (2922.28, 3610.35) | 1.02 (0.89, 1.18) | 69.39 (-399.67, 507.94) | 0.93 (0.90, 0.96) |
| Brock Model | 4732.08 (4325.85, 5184.41) | 5904.98 (5646.65, 6169.25) | 1.25 (1.15, 1.36) | 1172.89 (774.17, 1583.65) | 0.87 (0.85, 0.89) |

Abbreviations: CI, confidence interval.
[1] Our model was chosen based on K-fold cross-validations using the training sample, and performance metrics are reported based on the hold-out test sample. Our final model was a LASSO model.

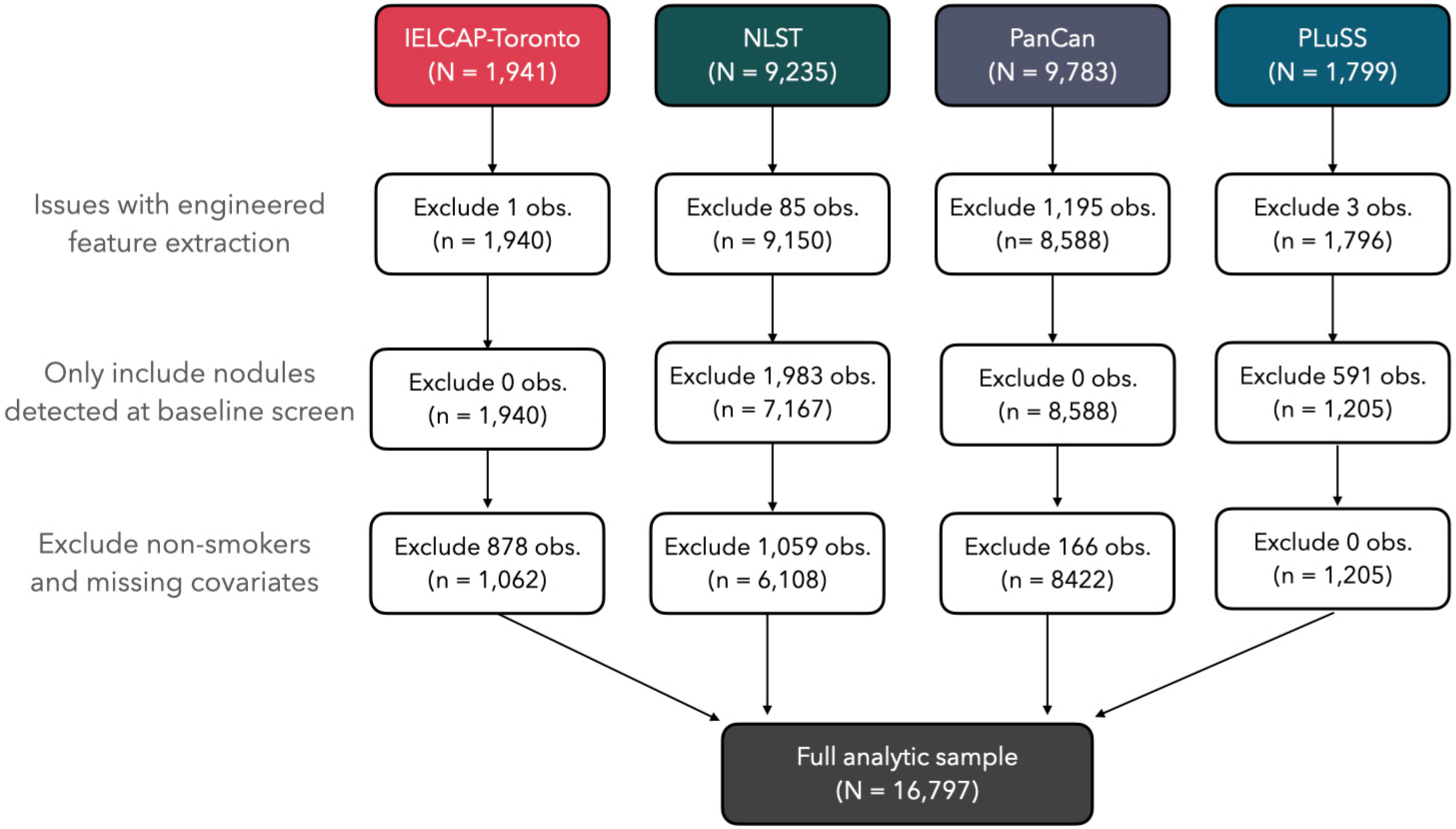


**Supplemental Figure 1.** Flow chart for the number of nodules retained for each of the four lung cancer screening studies based on sequential data cleaning steps to remove: (1) any nodules with feature extraction issues, (2) any nodules detected beyond the baseline CT scans, and (3) observations with missing patient-level epidemiologic data.

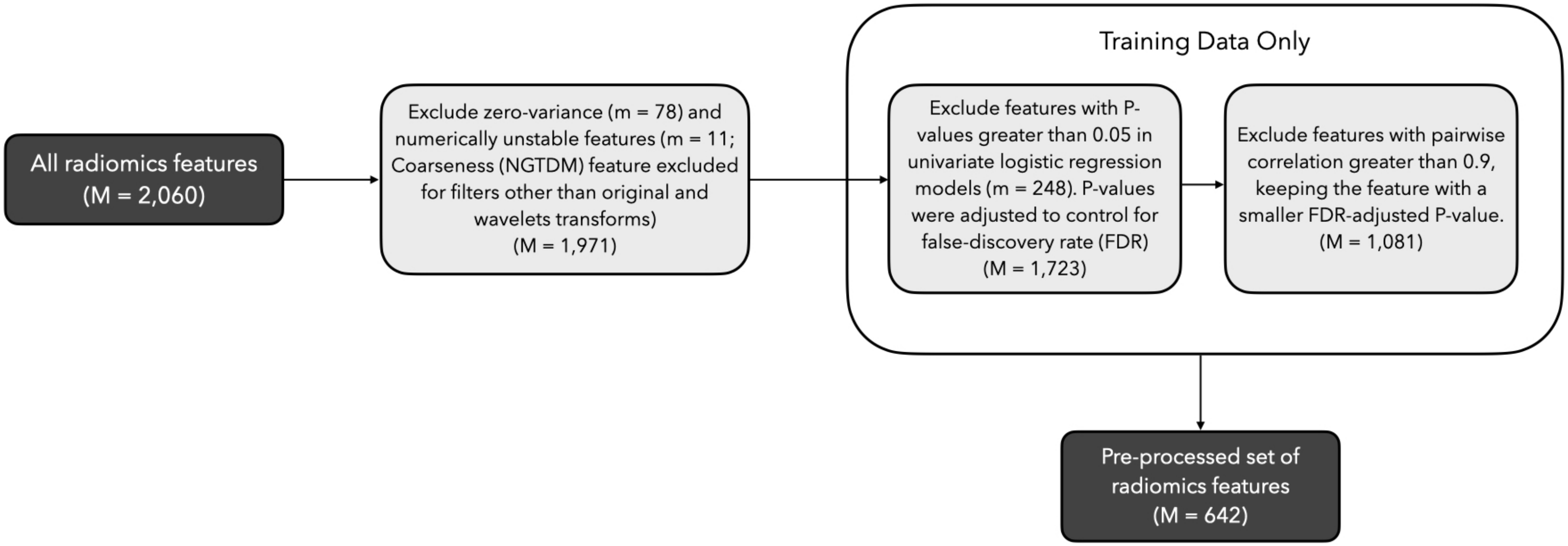


**Supplemental Figure 2.** Flow chart for the radiomics feature pre-processing steps to filter out (1) zero-variance and numerically unstable features, (2) non-significant features based on univariate p-values in the training data, and (3) highly correlated features in the training data, keeping the more significant feature.

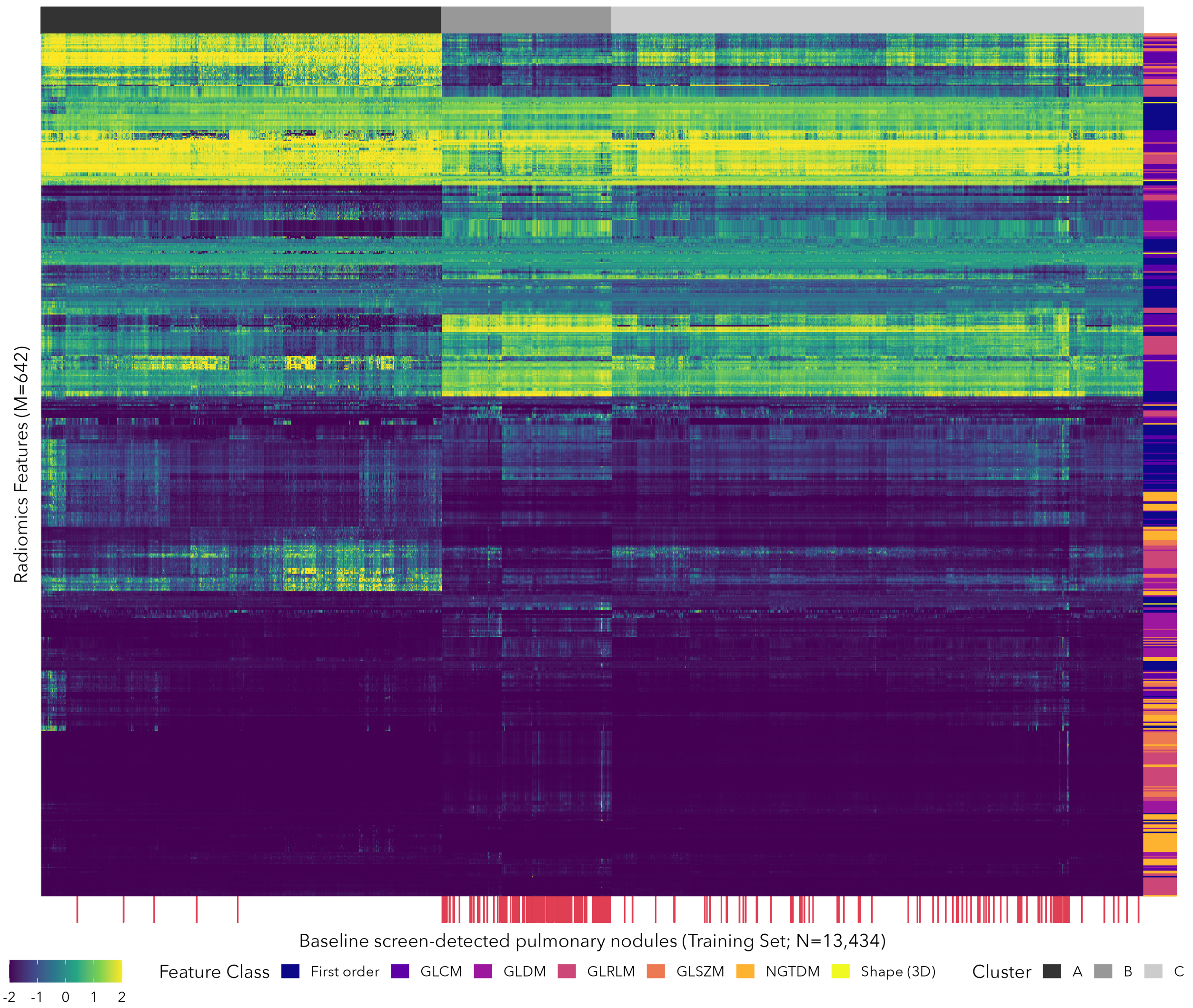


**Supplemental Figure 3.** Heat map for the 642 radiomics features retained in the feature filtering steps based on the training data (See Supplemental Figure 2). Features were min-max rescaled to the range [-2,2]. Three distinct clusters (i.e., radiomic profiles) were identified (indicated by the gray boxes along the top axis) and they differed significantly with respect to the proportion of malignant nodules (Fishers exact P-value < 0.05). Class membership for the 642 radiomic features are indicated along the right axis. Malignant nodules are indicated along the bottom axis.

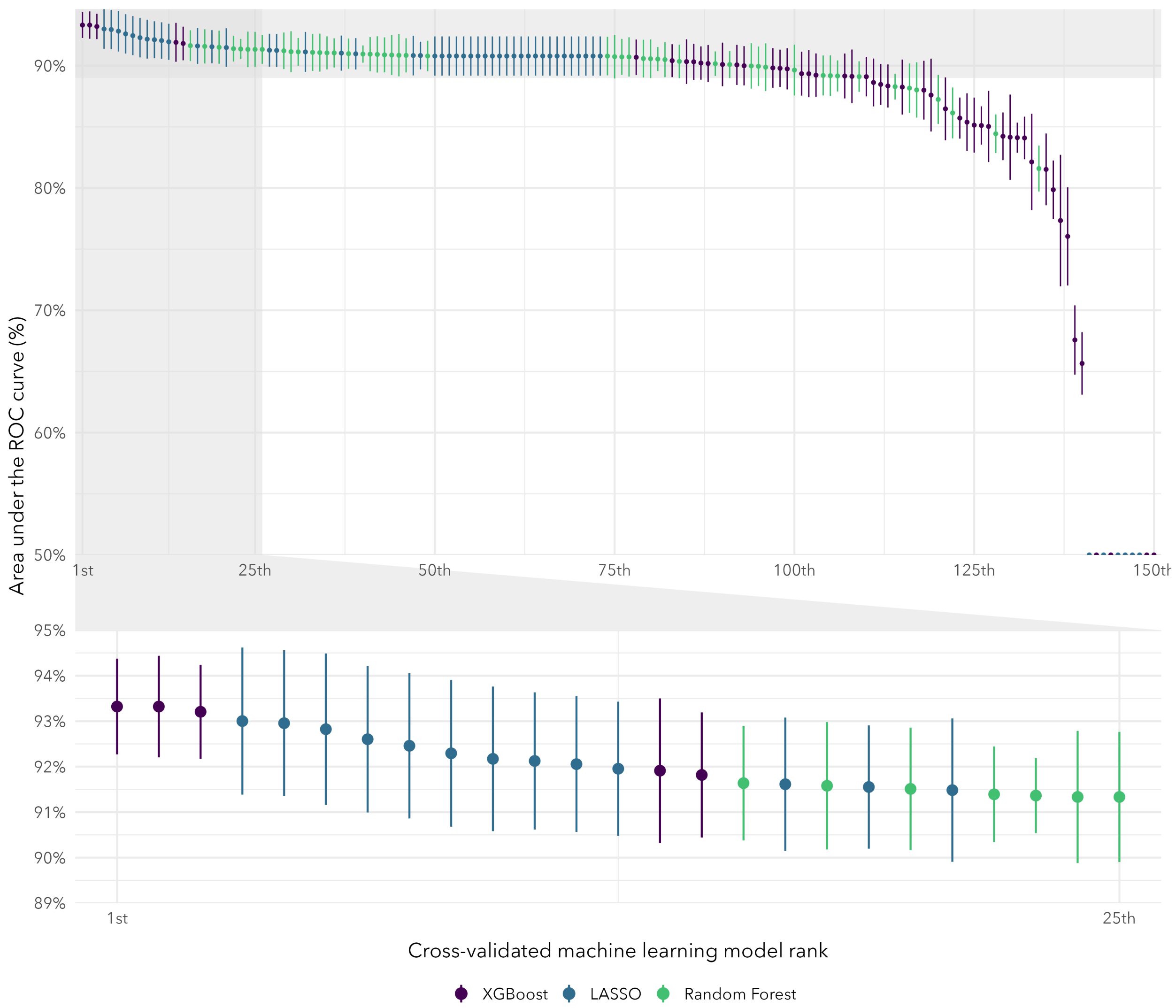


**Supplemental Figure 4.** Cross-validated area under the receiver operating characteristic curve (AUC) and 95% confidence intervals for each of the hyperparameter combinations (i.e., submodels) for the machine learning models. Models are ordered from left-to-right based on descending AUC. The top panel shows the rankings for every submodel and the bottom panel zooms in on the top 25 submodels.

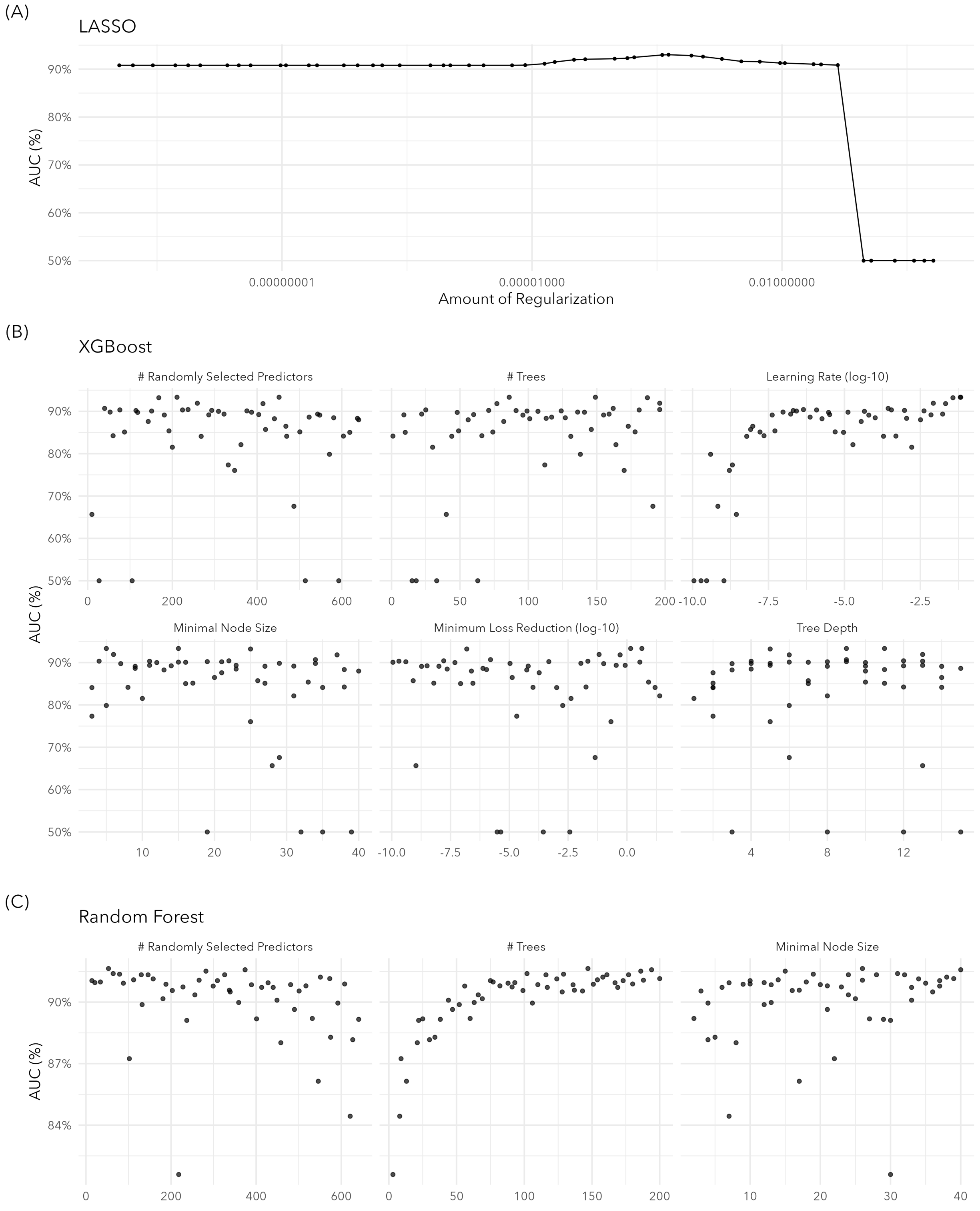


**Supplemental Figure 5.** Results from the five-fold cross-validation used to determine the top performing machine learning model. Area under the curve (AUC) are presented according to hyperparameters for (A) LASSO, (B) XGBoost, and (C) Random Forest.

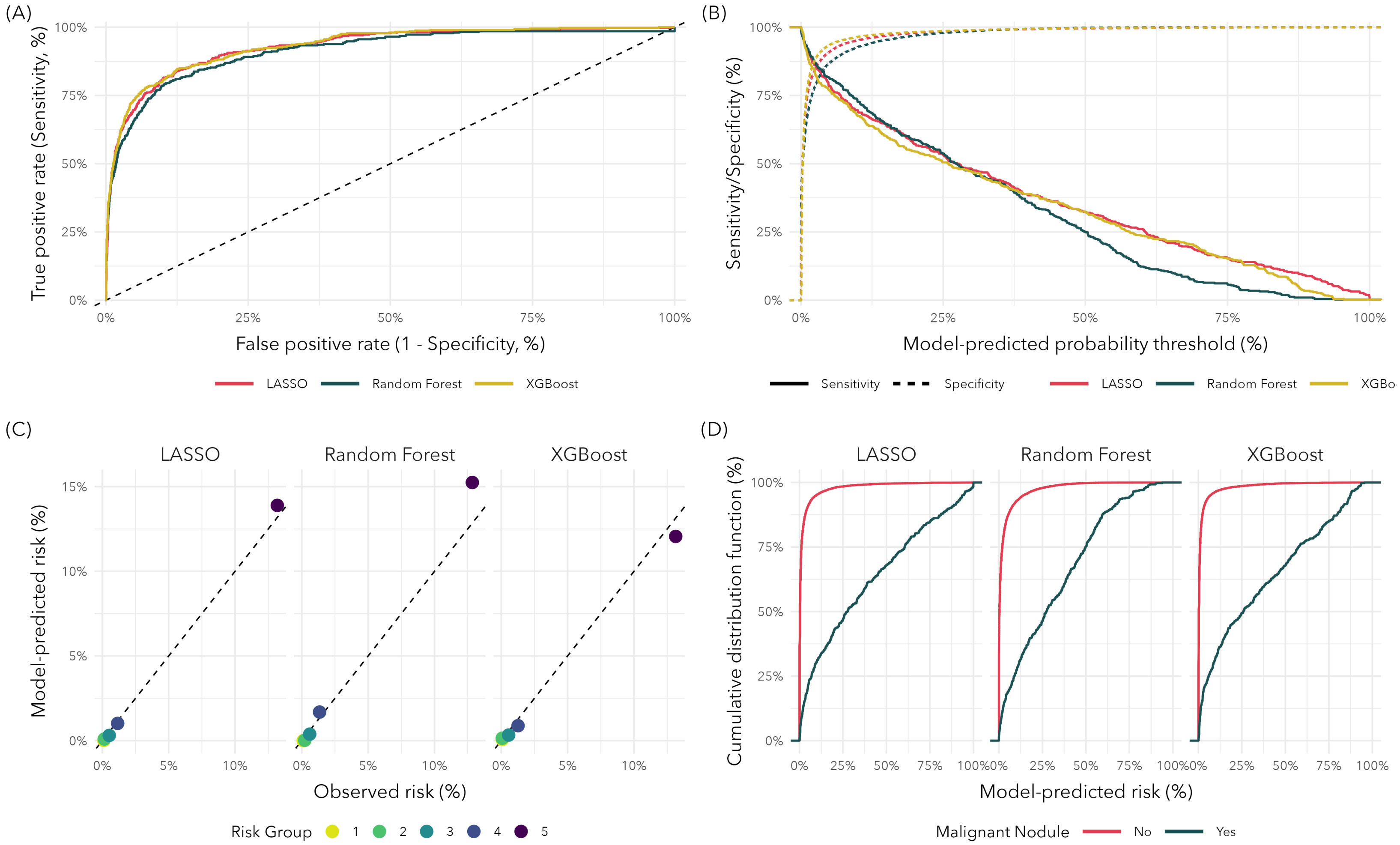


**Supplemental Figure 6.** Model performance metrics for the three top-performing machine learning models selected based on cross-validated area under the receiver operating characteristic curve (LASSO, Random Forest, XGBoost). (A) Receiver operating characteristic curves. (B) Sensitivity and specificity across the range of model-predicted probabilities. (C) Calibration plots comparing observed and model-predicted risks within quintiles of model-predicted risks. (D) Cumulative distribution functions of the model-predicted risks for benign and malignant pulmonary nodules.

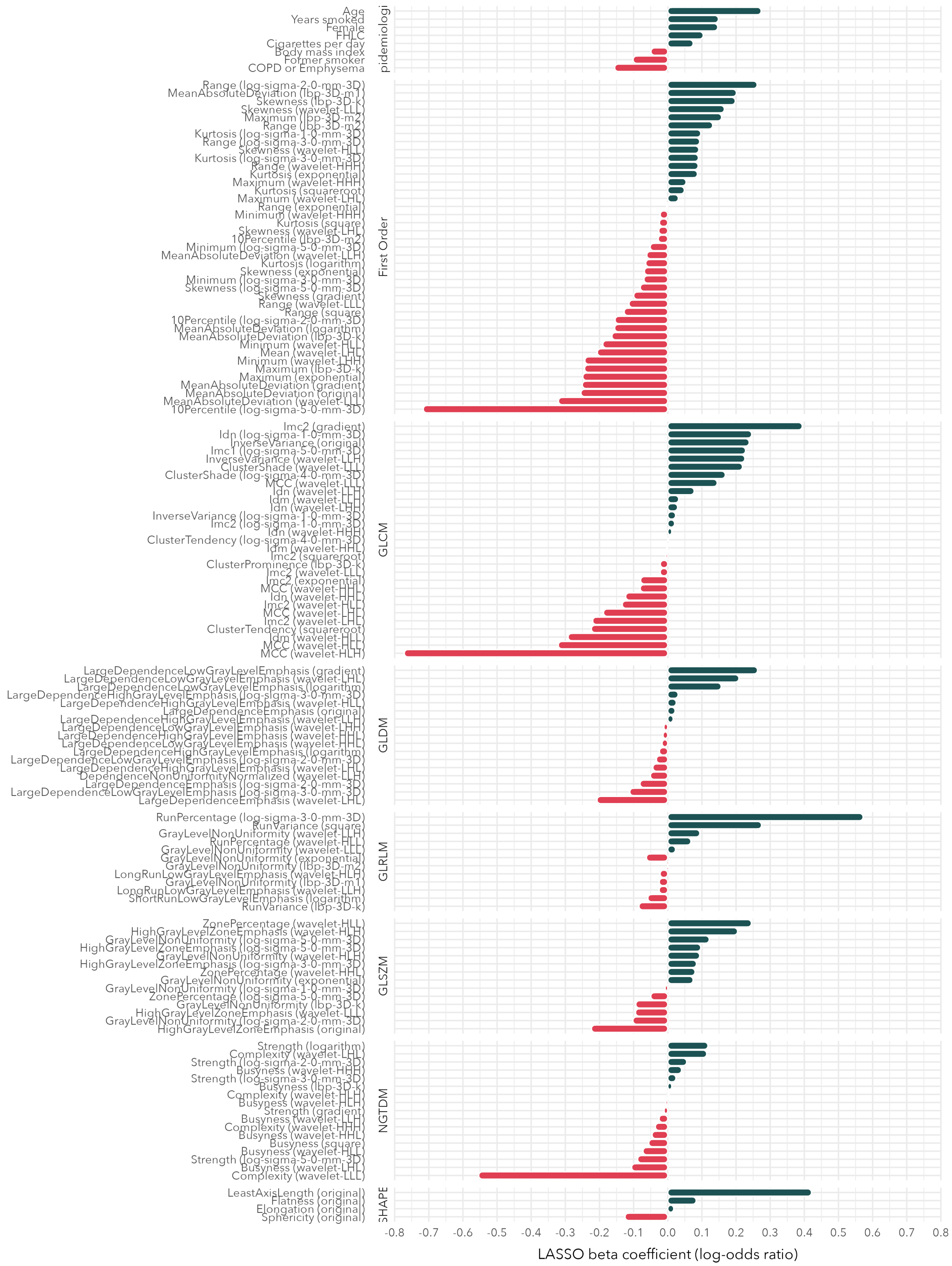


**Supplemental Figure 7.** Beta coefficients for the predictors retained in the final LASSO model with non-zero coefficients. Predictors are grouped by feature class. The image transformation associated with a radiomic feature is listed in parentheses.

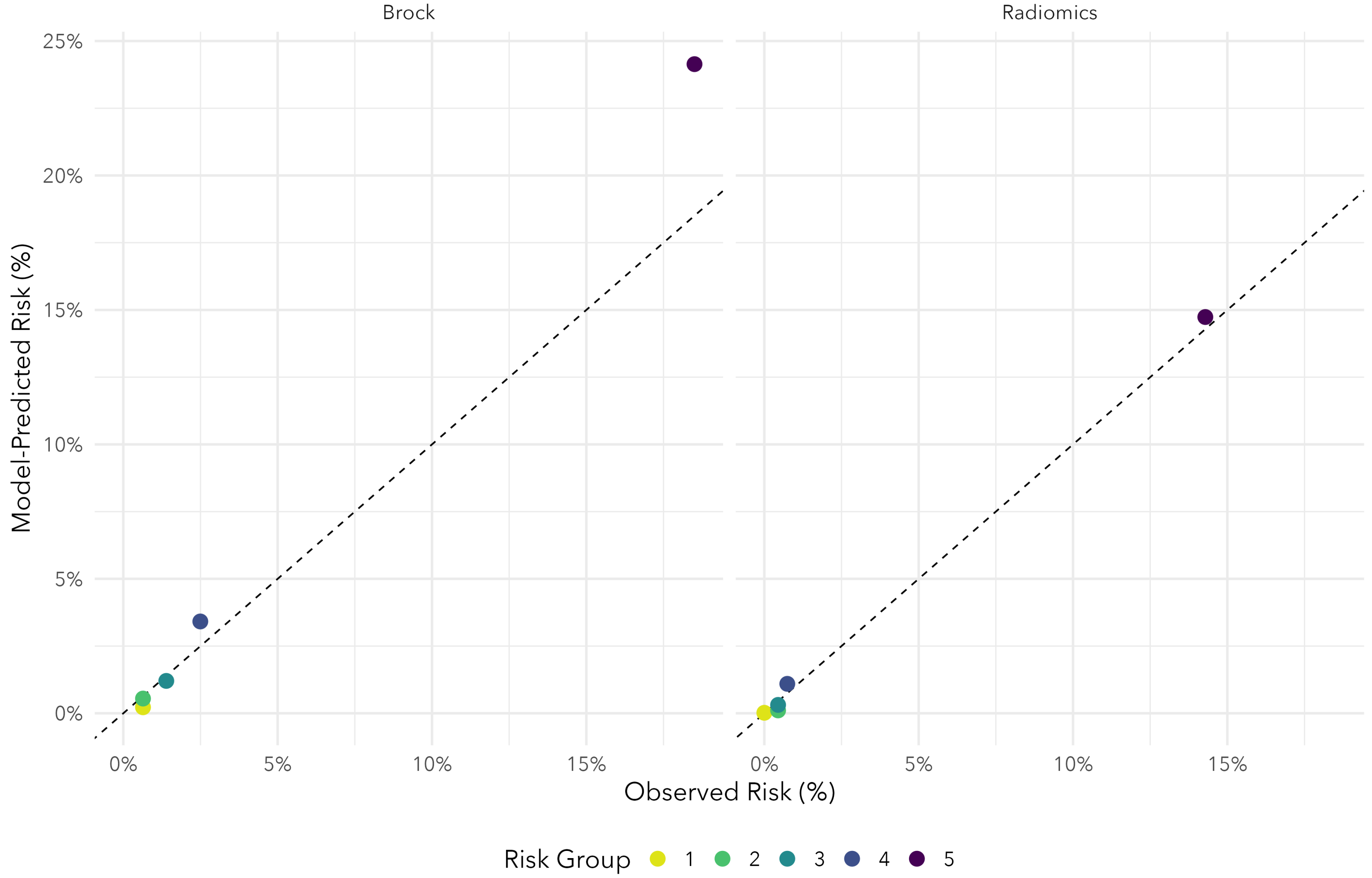


**Supplemental Figure 8.** Calibration plot comparing model-predicted risks to observed risks in the hold-out test-data for our radiomics model (LASSO) and the established Brock Model (McWilliams et al., 2013).
